# Epigenome-wide association studies of three social determinants of health and implications for lung functions among survivors of childhood cancer

**DOI:** 10.1101/2020.10.30.20223313

**Authors:** Nan Song, Jin-ah Sim, Qian Dong, Yinan Zheng, Lifang Hou, Zhenghong Li, Chia-Wei Hsu, Haitao Pan, Heather Mulder, John Easton, Emily Walker, Geoffrey Neale, Carmen L. Wilson, Kirsten K. Ness, Kevin R. Krull, Deo Kumar Srivastava, Yutaka Yasui, Jinghui Zhang, Melissa M. Hudson, Leslie L. Robison, I-Chan Huang, Zhaoming Wang

**Author notes:** N.S. and J.S. contributed equally as co-first authors. Z.W. and I-C.H. contributed equally as co-senior authors. **Correspondence:** Zhaoming Wang, Department of Epidemiology and Cancer Control, St. Jude Children’s Research Hospital, 262 Danny Thomas Place, MS 735, Memphis, TN 38105 USA.

## Abstract

**Background:** Emerging evidence suggests that social determinants of health (SDOH) may influence health and wellness through an epigenetic mechanism in the general population. However, the social epigenomic approach has not yet been applied to survivors of childhood cancer, a vulnerable population with elevated risk for chronic health conditions (CHCs).

**Methods:** Study participants were drawn from the St Jude Lifetime Cohort, a hospital-based retrospective cohort with prospective follow up. DNA methylation (DNAm) profiling was generated based on blood derived DNA collected during follow-up visit. SDOH included educational attainment, personal income, and area deprivation index (ADI) based on baseline or follow-up questionnaires and geocoding. CHCs were clinically assessed with severity grade.

**Results:** We included 258 childhood cancer survivors of African ancestry (AA) (median time from diagnosis=25.2 years, interquartile range [IQR]=19.9-32.1 years) and 1,618 survivors of European ancestry (EA) (median time from diagnosis=27.3, IQR=21.1-33.7 years). Through epigenome-wide association studies, we identified 130 SDOH-CpG associations including educational attainment (N=88), personal income (N=23), and ADI (N=19) at epigenome-wide significance level (*P*<9×10^−8^). There were 13 CpGs, commonly associated with all three SDOH factors, with attenuated remaining effect sizes (36.8-48.3%) after additionally adjusting body mass index and smoking, mapped to smoking-related genes including *GPR55, CLDND1, CPOX, GPR15, AHRR, PRRC2B*, and *ELMSAN1*. Among 130 SDOH-related CpGs, three independent CpGs (cg04180924, cg1120500, and cg27470486) had a significant combined mediation effect for educational attainment (%mediation=48.9%), and a single mediator cg08064403 was found with significant mediation effect for personal income (25.9%) and ADI (24.1%) on pulmonary diffusion deficit, which showed higher incidence in AA than in EA survivors implying racial disparity which is possibly due to more disadvantageous SDOH factors in AA than in EA.

**Conclusions:** We demonstrated striking DNAm signatures associated with multiple SDOH factors (educational attainment, personal income, and ADI) and many epigenome-wide significant CpG sites resembling the effect of smoking exposure. We also identified an exemplified racial health disparity in pulmonary diffusion deficit between AA and EA survivors and illuminated DNAm as potential mechanistic mediators for SDOH factors using a social epigenomic approach.

## Introduction

Previous research suggested that childhood cancer survivors of African ancestry (AA) had significantly poorer survivorship including higher mortality and prevalent cardiovascular disorders, stroke, diabetes, hypertension, and neurocognitive and emotional outcomes compared to non-Hispanic Caucasian survivors.^1-4^ However, after adjusting for the influence of personal socio-economic factors, health disparities were either substantially decreased or became not statistically significant,^1-4^ suggesting the importance of socio-economic factors in racial health disparity among survivors of childhood cancer.

Social and behavioral epigenomics is an emerging transdisciplinary field studying social and the relevant health behavioral factors that can influence health outcomes through an epigenetic mechanism and how they may contribute to health disparities.^5,6^ Among a variety of epigenetic modifications, DNA methylation (DNAm) has been the most widely studied, reporting significant associations with overall socio-economic status (SES),^5,7-11^ educational attainment,^12,13^ and health behaviors.^14^ The epigenetic signatures may further link SES to chronic disease risk.^10^ Recently, we reported the large-scale epigenetic study, showing that cancer treatment exposures were associated with persistent variations of methylation in DNA derived from blood decades later and a subset of treatment-associated DNAm 5’-cytosine-phosphate-guanine-3’ (CpG) sites was also significantly associated with cardiometabolic risk.^15^ Although intensive treatment exposures would be expected to leave persistent epigenetic marks and shape up the survivors’ methylome, poor social and behavioral factors during the life-course may pose considerable and enduring impact on DNAm among aging survivors.

In this study, we hypothesize that DNAm is a mechanism for biological embedding of life experiences including social determinants of health (SDOH) and associated health behaviors. SDOH-related DNAm variations may mediate the association between poor SDOH factors and elevated risk of chronic health conditions (CHC). It has been suggested that epigenetic marks, such as DNAm, could influence the transcription potential of genomic regions and, once changed, could have long-term effects.^16^ Furthermore, health behaviors and other environmental factors, such as body mass index (BMI) and smoking which were known to be associated with CHCs, might have a role in this pathway. Systematic epigenome-wide association studies (EWAS) investigating the associations among SDOH factors, DNAm, and CHCs later in life have not yet been conducted in survivors of childhood cancer, even though survivors have much higher burden of morbidity and mortality and tend to be more sensitive to social stressors compared to the general population. To fill this gap, we employed an epigenome-wide approach to systematically identify the DNAm CpGs associated with three key SDOH factors, namely, educational attainment, personal income, and neighborhood SES and physical environment, and evaluated whether SDOH-associated CpGs are also associated with a focused set of pulmonary conditions, related to lung function, which were potentially relevant to smoking. We further applied mediation analysis to explore potential role of DNAm in mediating the effect of each SDOH factor on risk of these conditions.

## Results

### Characteristics of study population

Study participants included childhood cancer survivors of AA (n=258, median time from diagnosis=25.2 years, interquartile range [IQR]=19.9-32.1 years) and survivors of European ancestry (EA) (n=1,618, median time from diagnosis=27.3 years, IQR=21.1-33.7 years) (Table 1). The median age at primary diagnosis was 9.6 (IQR=4.2-14.4) years in AA and 9.0 (IQR=3.8-14.4) years in EA and median age at DNA sampling was 33.9 (IQR=29.4-39.6) years in AA and 35.3 (IQR=30.0-42.1) years in EA. The proportion of female was 53.1% in AA and 40.4% in EA. More than 60% (61.6% in AA and 61.7% in EA) was overweight (≥25kg/m^2^). Smoking was significantly different with 70.2% of non-smokers in AA and 58.2% of non-smokers in EA (*P*<0.0001). Three SDOH factors were significantly different between AA and EA; specifically, AA had the lower educational attainment (*P*<0.0001), lower personal income (*P*<0.0001), and lived in neighborhoods with poorer socio-economic and physical environment measured by the area deprivation index (ADI) (*P*<0.0001) than EA. Primary cancer diagnoses and cancer treatment information for the study population are also listed in **Table 1**.

**Table 1.**
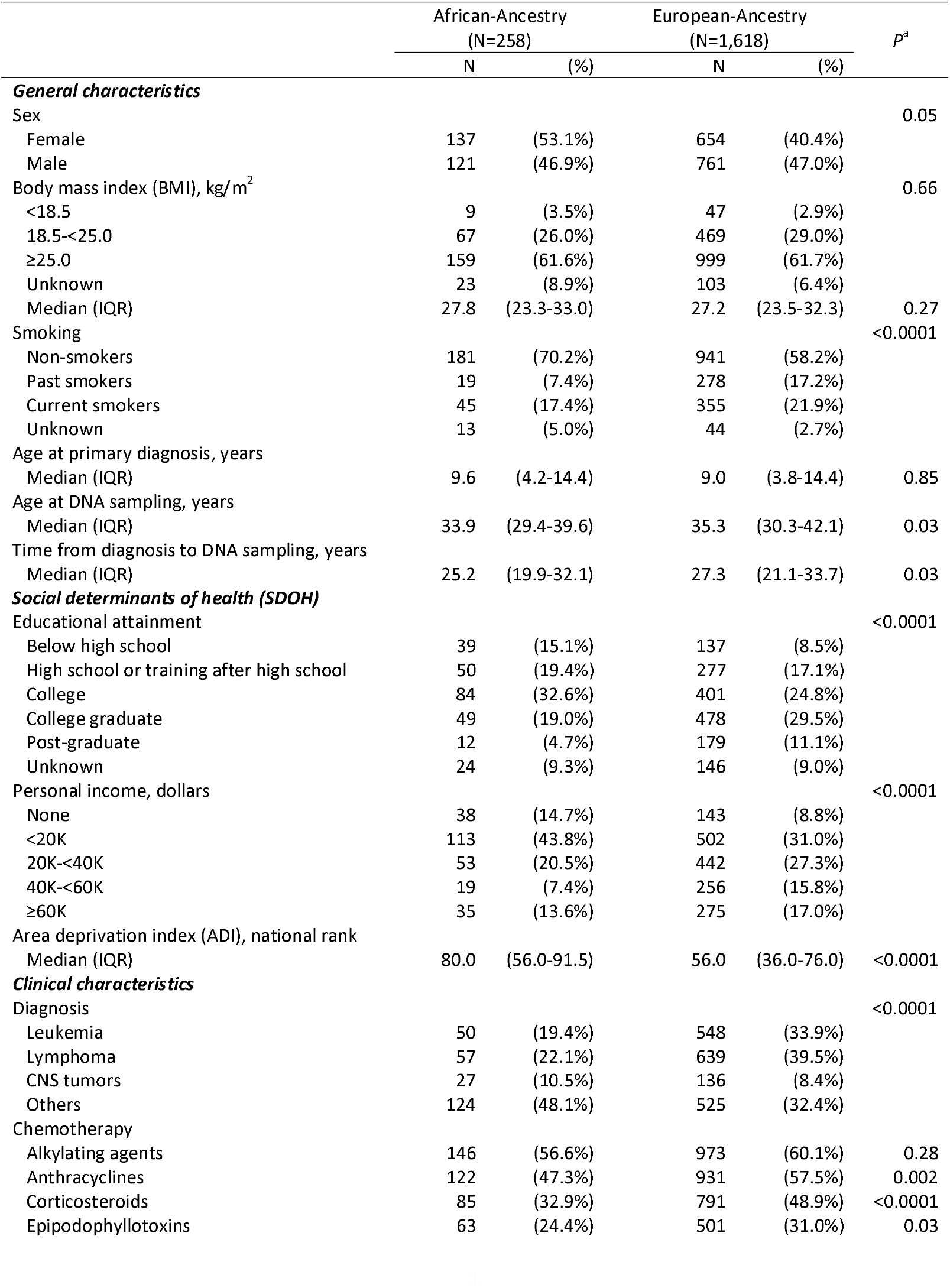
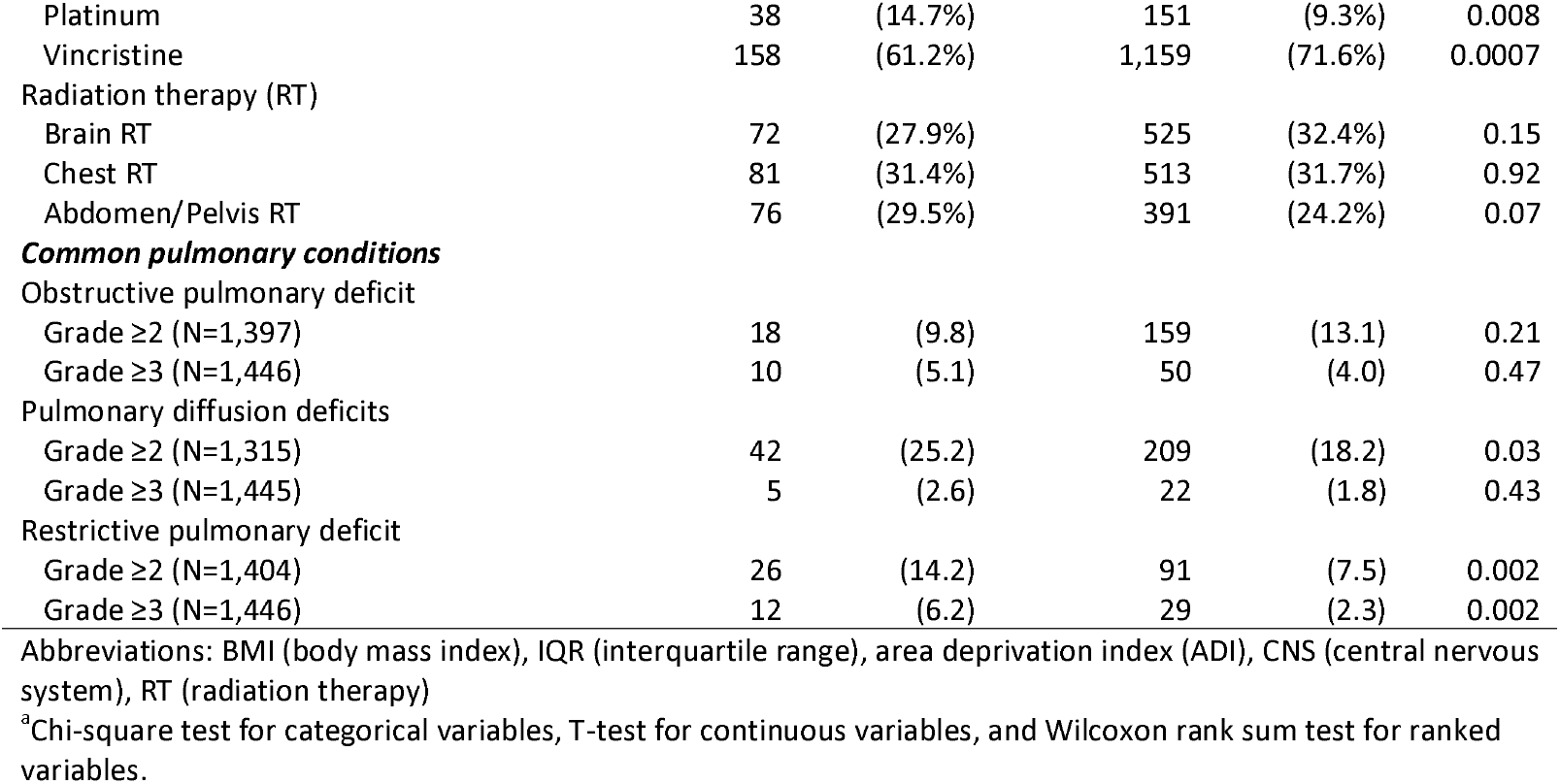
Characteristics of study participants

|  | African-Ancestry<br>(N=258) |  | European-Ancestry<br>(N=1,618) |  | P<sup>a</sup> |
| --- | --- | --- | --- | --- | --- |
|  | N | (%) | N | (%) |  |
| **General characteristics** |  |  |  |  |  |
| Sex |  |  |  |  | 0.05 |
| Female | 137 | (53.1%) | 654 | (40.4%) |  |
| Male | 121 | (46.9%) | 761 | (47.0%) |  |
| Body mass index (BMI), kg/m<sup>2</sup> |  |  |  |  | 0.66 |
| <18.5 | 9 | (3.5%) | 47 | (2.9%) |  |
| 18.5-<25.0 | 67 | (26.0%) | 469 | (29.0%) |  |
| ≥25.0 | 159 | (61.6%) | 999 | (61.7%) |  |
| Unknown | 23 | (8.9%) | 103 | (6.4%) |  |
| Median (IQR) | 27.8 | (23.3-33.0) | 27.2 | (23.5-32.3) | 0.27 |
| Smoking |  |  |  |  | <0.0001 |
| Non-smokers | 181 | (70.2%) | 941 | (58.2%) |  |
| Past smokers | 19 | (7.4%) | 278 | (17.2%) |  |
| Current smokers | 45 | (17.4%) | 355 | (21.9%) |  |
| Unknown | 13 | (5.0%) | 44 | (2.7%) |  |
| Age at primary diagnosis, years |  |  |  |  |  |
| Median (IQR) | 9.6 | (4.2-14.4) | 9.0 | (3.8-14.4) | 0.85 |
| Age at DNA sampling, years |  |  |  |  |  |
| Median (IQR) | 33.9 | (29.4-39.6) | 35.3 | (30.3-42.1) | 0.03 |
| Time from diagnosis to DNA sampling, years |  |  |  |  |  |
| Median (IQR) | 25.2 | (19.9-32.1) | 27.3 | (21.1-33.7) | 0.03 |
| **Social determinants of health (SDOH)** |  |  |  |  |  |
| Educational attainment |  |  |  |  | <0.0001 |
| Below high school | 39 | (15.1%) | 137 | (8.5%) |  |
| High school or training after high school | 50 | (19.4%) | 277 | (17.1%) |  |
| College | 84 | (32.6%) | 401 | (24.8%) |  |
| College graduate | 49 | (19.0%) | 478 | (29.5%) |  |
| Post-graduate | 12 | (4.7%) | 179 | (11.1%) |  |
| Unknown | 24 | (9.3%) | 146 | (9.0%) |  |
| Personal income, dollars |  |  |  |  | <0.0001 |
| None | 38 | (14.7%) | 143 | (8.8%) |  |
| <20K | 113 | (43.8%) | 502 | (31.0%) |  |
| 20K-<40K | 53 | (20.5%) | 442 | (27.3%) |  |
| 40K-<60K | 19 | (7.4%) | 256 | (15.8%) |  |
| ≥60K | 35 | (13.6%) | 275 | (17.0%) |  |
| Area deprivation index (ADI), national rank |  |  |  |  |  |
| Median (IQR) | 80.0 | (56.0-91.5) | 56.0 | (36.0-76.0) | <0.0001 |
| **Clinical characteristics** |  |  |  |  |  |
| Diagnosis |  |  |  |  | <0.0001 |
| Leukemia | 50 | (19.4%) | 548 | (33.9%) |  |
| Lymphoma | 57 | (22.1%) | 639 | (39.5%) |  |
| CNS tumors | 27 | (10.5%) | 136 | (8.4%) |  |
| Others | 124 | (48.1%) | 525 | (32.4%) |  |
| Chemotherapy |  |  |  |  |  |
| Alkylating agents | 146 | (56.6%) | 973 | (60.1%) | 0.28 |
| Anthracyclines | 122 | (47.3%) | 931 | (57.5%) | 0.002 |
| Corticosteroids | 85 | (32.9%) | 791 | (48.9%) | <0.0001 |
| Epipodophyllotoxins | 63 | (24.4%) | 501 | (31.0%) | 0.03 |
| Platinum | 38 | (14.7%) | 151 | (9.3%) | 0.008 |
| Vincristine | 158 | (61.2%) | 1,159 | (71.6%) | 0.0007 |
| Radiation therapy (RT) |  |  |  |  |  |
| Brain RT | 72 | (27.9%) | 525 | (32.4%) | 0.15 |
| Chest RT | 81 | (31.4%) | 513 | (31.7%) | 0.92 |
| Abdomen/Pelvis RT | 76 | (29.5%) | 391 | (24.2%) | 0.07 |
| ***Common pulmonary conditions*** |  |  |  |  |  |
| Obstructive pulmonary deficit |  |  |  |  |  |
| Grade ≥2 (N=1,397) | 18 | (9.8) | 159 | (13.1) | 0.21 |
| Grade ≥3 (N=1,446) | 10 | (5.1) | 50 | (4.0) | 0.47 |
| Pulmonary diffusion deficits |  |  |  |  |  |
| Grade ≥2 (N=1,315) | 42 | (25.2) | 209 | (18.2) | 0.03 |
| Grade ≥3 (N=1,445) | 5 | (2.6) | 22 | (1.8) | 0.43 |
| Restrictive pulmonary deficit |  |  |  |  |  |
| Grade ≥2 (N=1,404) | 26 | (14.2) | 91 | (7.5) | 0.002 |
| Grade ≥3 (N=1,446) | 12 | (6.2) | 29 | (2.3) | 0.002 |

Abbreviations: BMI (body mass index), IQR (interquartile range), area deprivation index (ADI), CNS (central nervous system), RT (radiation therapy)
<sup>a</sup>Chi-square test for categorical variables, T-test for continuous variables, and Wilcoxon rank sum test for ranked variables.

### Association of DNA methylation sites with SDOH factors

First, EWAS was conducted in EA. The association P-values in -log_10_ scale were shown for three series of EWAS analyses across the genome in the circos plot (**Figure 1**), with 337 for educational attainment, 39 for personal income, and 24 CpG sites for ADI reached over epigenome-wide significance level with *P*<9×10^−8^, respectively. There were 13 significant DNAm CpGs commonly associated with all three SDOH factors. Those 13 CpGs were mapped to the five genomic regions on chromosomes 2, 3, 5, 9, and 14 harboring genes including *GPR55, CLDND1, CPOX, GPR15, AHRR, PRRC2B*, and *ELMSAN1*, which are previously known to be associated with smoking exposures (Table 2). After additionally adjusting for BMI and smoking, all 13 CpGs remained statistically significant (*P*<0.05), but with attenuated effect sizes remaining on average 36.8% for educational attainment (range=30.8%-48.8%), 48.3% for personal income (range=38.5-58.8%), and 43.3% for ADI (range=35.1-61.5%). Given SDOH-CpG associations related to and attenuated by smoking, this finding prompted us to hypothesize that health outcomes related to lung functions may be most likely impacted by the SDOH-associated CpGs.

**Table 2.**
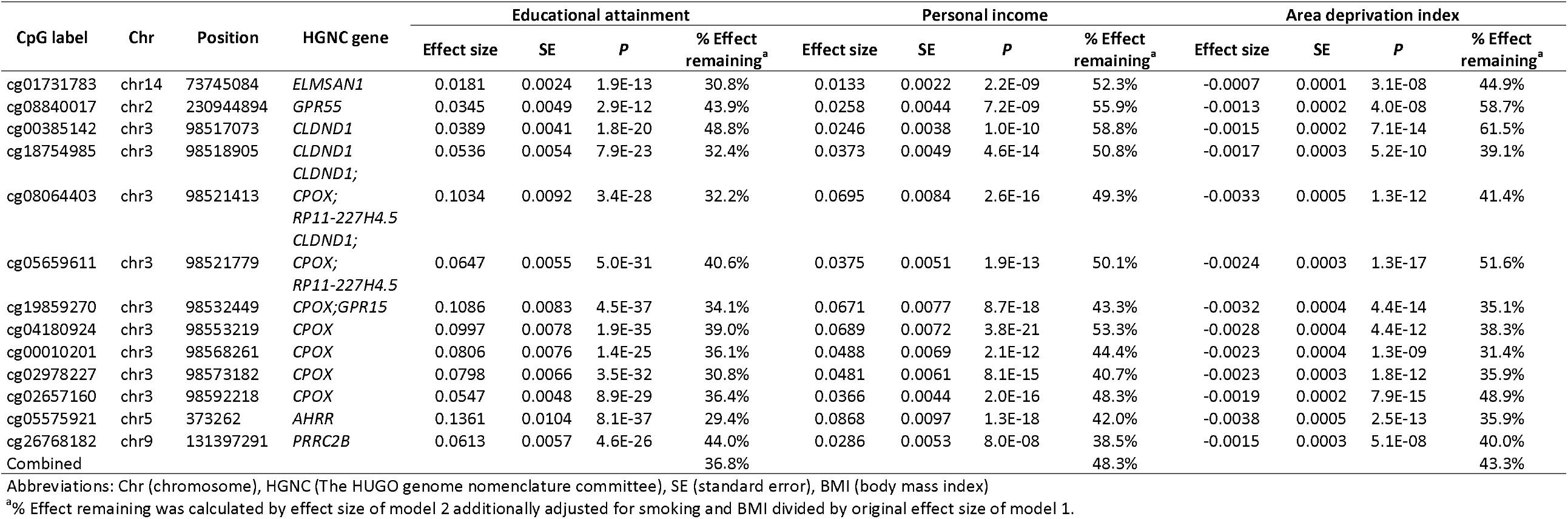
DNA methylation CpG sites that are significantly associated with all three social determinants of health factors

**Figure 1.**
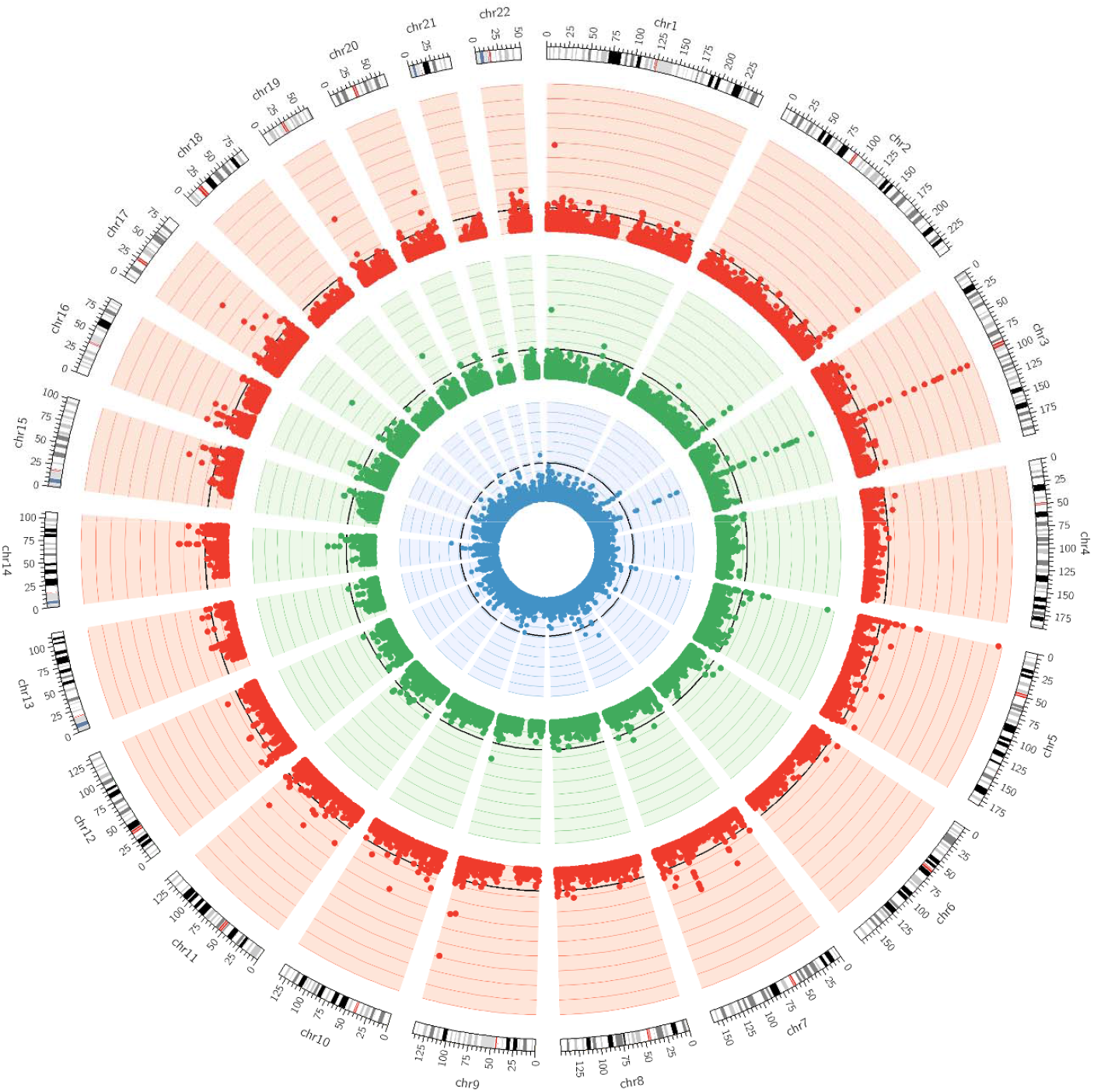
Circos plot for epigenome-wide association studies (EWAS) of three social determinants of health factors. Outer circle (red): EWAS for educational attainment; middle circle (green): EWAS for personal income; inner circle (blue): EWAS for area deprivation index. Each dot depicts -log10 p value for each DNAm CpG site mapped to a chromosome location along the genome. The black lines indicate the epigenome-wide significance level (*P*=9×10^−8^).

In the EA model additionally adjusted by BMI and smoking, there were still three epigenome-wide significant associations between educational attainment and cg04180924 (chr3, CPOX, *P*=2.0×10^−8^), cg04885881 (chr1, intergenic, *P*=1.3×10^−9^), cg06359375 (chr22, *HPS4*, P=2.0×10^−8^). There was one epigenome-wide significant association between personal income and cg04180924 (chr3, *CPOX, P*=5.4×10^−9^), but no single CpG reached epigenome-wide significant level for ADI. We chose to report the models without adjusting for BMI and smoking as the primary results, however, we applied the condition of *P*<0.05 in the model additionally adjusted by BMI and smoking to filter against all epigenome-wide significant DNAm CpG sites in the corresponding models without BMI and smoking adjustments. Following this filtering step, we had 130 SDOH-CpG associations including educational attainment (N=88), personal income (N=23), and ADI (N=19). The detailed estimates were provided (**Supplementary Table 1**).

Similarly, EWAS analyses were performed in AA survivors and none of DNAm CpGs reached epigenome-wide significance level (*P*<9×10^−8^). However, for comparison purpose, we presented the AA results for 130 SDOH-CpG associations statistically established in EA survivors **(Supplementary Table 2**). Among 130 SDOH-CpG associations, 25 associations were also validated in AA with *P*<0.05; specifically, 15 for educational attainments, 8 for personal income, and 3 for ADI.

### Association of social determinants of health with pulmonary conditions

The incidence of three common pulmonary conditions were examined among EA and AA survivors using 2 or 3 as the minimum Common Terminology Criteria for Adverse Events (CTCAE)-grade (**Table 1 and Figure 2**). In the unadjusted model with minimum CTCAE-grade set to 2, pulmonary diffusion deficit was significantly higher in AA (25.2% in AA and 18.2% in EA, *P*=0.03), and restrictive pulmonary deficit was also significantly higher in AA (14.2% in AA and 7.5% in EA, P=0.002), whereas obstructive pulmonary deficit was comparable between the two racial groups (9.8% in AA and 13.1% in EA, P=0.21). In the model adjusting for other covariates, regardless of with or without SDOH factors, race was significantly associated with restrictive pulmonary deficits. However, race was significantly associated with pulmonary diffusion deficits only in the model without adjusting for SDOH factors and became not significant after adjusting for SDOH (**Supplementary Table 3**), suggesting significant contribution of SDOH factors to association between race and pulmonary diffusion deficits.

**Table 3.** DNA methylation partially mediates the association between social determinants of health factors and risk of pulmonary diffusion deficits

| Social determinants of Health | CpG | Chromosome | Position | HGNC gene | ACME | ADE | Total effect<sup>a</sup> | % Mediation<sup>b</sup> | $P_{FDR}$ |
| --- | --- | --- | --- | --- | --- | --- | --- | --- | --- |
| Educational attainment | cg04180924 | chr3 | 98,553,219 | *CPOX* | -0.0137 | -0.0280 | -0.0417 | 32.9% | 0.01 |
|  | cg11205006 | chr22 | 26,479,532 | *HPS4* | -0.0058 | -0.0246 | -0.0304 | 19.0% | 0.02 |
|  | cg27470486 | chr17 | 41,917,434 | *ACLY* | -0.0029 | -0.0307 | -0.0336 | 8.6% | 0.04 |
|  | Combined Score<sup>a</sup> |  |  |  | -0.0214 | -0.0224 | -0.0438 | 48.9% | 0.002 |
| Personal income | cg08064403 | chr3 | 98,521,413 | *CLDND1;CPOX;RP11-227H4.5* | -0.0101 | -0.0290 | -0.0392 | 25.9% | <0.001 |
| Area deprivation index | cg08064403 | chr3 | 98,521,413 | *CLDND1;CPOX;RP11-227H4.5* | 0.0002 | 0.0007 | 0.0009 | 24.1% | <0.001 |

Abbreviations: Chr (chromosome), HGNC (The HUGO genome nomenclature committee), ACME (average causal mediation effect), ADE (average direct effect), FDR (false-discovery rate)
<sup>a</sup>Calculated by summing ACME and ADE.
<sup>b</sup>Calculated by ACME divided by total effect (the sum of ACME and ADE).

**Figure 2.**
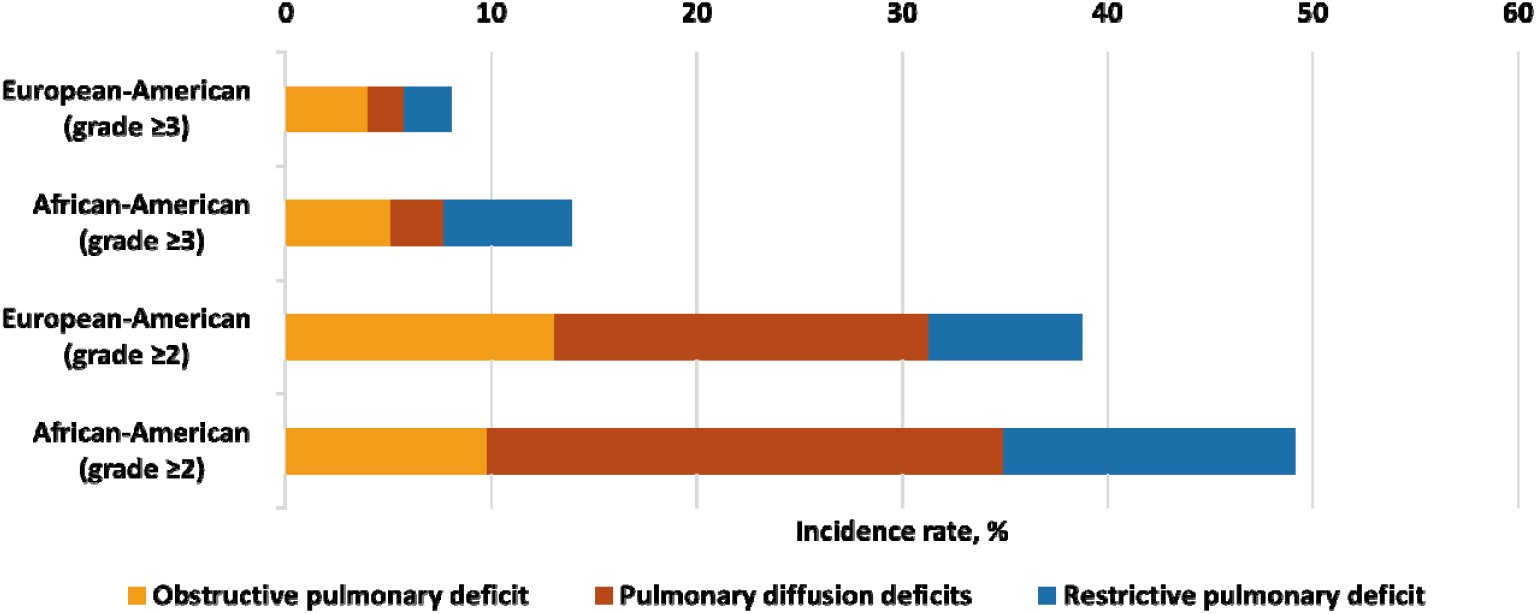
Incidence of common pulmonary conditions among survivors of African ancestry and European ancestry

### Methylation mediation effect for social determinants of health on pulmonary conditions

Among all 130 SDOH-CpG associations from EWAS in EA survivors, 61 CpGs (29 educational attainment associated CpG sites, 16 personal income associated CpGs sites, and 16 ADI associated CpG sites) were also significantly associated with risk of pulmonary diffusion deficits after adjusting for multiple comparisons (*P*_FDR_<0.05) (**Supplementary Table 4**). In the mediation analysis, 17 out of 29 educational attainment associated CpGs were identified with significant average causal mediation effects (ACME) after adjusting for multiple comparisons. Using squared pairwise Pearson correlation coefficient r^2^ threshold of 0.05, three independent CpGs including cg04180924 (chr3, CPOX, mediation=32.9%, P_FDR_=0.01), cg11205006 (chr3, CPOX, mediation=32.9%, P_FDR_=0.01), and cg27470486 (chr17, ACLY, mediation=8.6%, P_FDR_=0.04) were obtained by top-down pruning the 17 CpGs sorted by estimated ACME in decreasing order. For the final mediation analysis, a combined score (i.e., summation of DNAm levels of the three CpGs with the same direction of association) was used as the mediating variable and 48.9% of mediation effect for educational attainment on pulmonary diffusion deficits was achieved (Table 3). Similarly, single significant mediator cg08064403 was found with significant partial mediation effect for personal income (mediation=25.9%, P<0.001) and ADI (mediation=24.1%, P<0.001) on pulmonary diffusion deficits (**Table 3**).

## Methods

### Study population

The St. Jude Lifetime Cohort (SJLIFE) study, a retrospective hospital-based cohort study for survivors of childhood cancer with prospective clinical follow-up, has been described elsewhere.^17,18^ Survivors completed demographic and epidemiological questionnaires, received clinical assessments at follow-up visits. A total of 2,052 EA survivors and 370 AA survivors also had DNAm profiling data generated with EPIC array.^15^ We subsequently excluded survivors with attained age younger than 25 years old (n=533) to ensure most individuals should have possibly attained the highest level of education (i.e., post-college), and principal components analysis (PCA) outliers (n=13). The remaining 1,618 EA and 258 AA survivors were included in this current study.

### Social determinants of health

SDOH factors for cancer survivors at personal and neighborhood levels were considered and collected at the same time as or before DNA sampling. The personal level factors included educational attainment and personal income measures. Neighborhood social determinants included area SES deprivation as a risk factor. Educational attainment and personal income were collected from SJLIFE surveys. Personal educational attainment was categorized into five levels (below high school, high school or training after high school, some college, college graduate, and post-graduate), and household incomes was categorized into five levels (None, <20, 20K to <40K, 40 to <60K, and ≥60K). We evaluated area deprivation of each survivor using the ADI that is comprised of 17 neighborhood-based SES measures including income, employment, education, and housing status collected in American Community Survey.^19,20^ Census track and block level of data were created by geocoding the participants’ residential addresses using Geographic Identifiers (GEOIDs) from Census Bureau, then geocoded data was linked to national ADI file. Technically, these measures present SES and physical environment by census blocks in the USA, and each census block receives a percentile ranking with minimum disadvantage in the 1st percentile and maximum disadvantage in the 100th percentile. For ADI, we considered >75th percentile, 40 to 75 percentile, and <40 percentile as high, moderate, and low area SES deprivation, respectively.

### Chronic health conditions

We will focus on the following three common CHC related to lung function: obstructive pulmonary deficit, pulmonary difficult deficit, and restrictive pulmonary deficit. Clinical evaluations of these common conditions have been documented in previous publications for SJLIFE cohort.^17,18^ CHCs with severity grading based on CTCAE version 4.0: 0 (no problem), 1 (mild), 2 (moderate), 3 (severe/disabling), 4 (life-threatening), or 5 (death).^18^ Only incident CHC with grade ≥2 and occurred after blood draw for DNAm profiling were considered as an adverse event in the current study.

### DNA methylation profiling

Blood samples collected from each participant at their follow-up visits were banked in the St. Jude Biorepository. Methylation profiling were performed using the Infinium® MethylationEPIC BeadChip. Genomic DNA (250ng per sample) were treated with bisulfite using the Zymo EZ DNA Methylation Kit under the following thermos-cycling conditions: 16 cycles: 95°C for 30 sec, 50°C for 1 hour. Following bisulfite treatment, DNA samples were desulphonated, column purified, then eluted using 12 ul of elution buffer (Zymo Research Corp.). Treating DNA with sodium bisulfite selectively converted cytosine to uracil but preserves 5-methylcytosine, which was immune from deamination. Bisulfite-converted DNA (4 µl) was then processed by following the Illumina Infinium Methylation Assay protocol, which included hybridization to MethylationEPIC BeadChips, single base extension assay, and staining and scanning using the Illumina HiScan system.

### DNA methylation bioinformatic analysis

Methylation raw intensity data were analyzed in R (version 3.6.1) using minfi package.^21^ The processed methylation data was described as a β-value, which is a continuous variable ranging between 0 (no methylation) and 1 (full methylation). In any sample, a probe with a detection p value more than 0.01 was assigned missing status. Any sample or probe with more than 5% missing values were excluded from downstream analysis. Non-specific or cross-reactive probes or probes with SNPs nearby the CpG site were also be excluded. The methylation data were normalized by quantile normalization method implemented in minfi package.^21^ M-value (logit transformation of β-value) were calculated and subsequently used in regression analyses.^22^ The epigenome-wide methylation scores were used to impute six lymphocyte cell subtype proportions (lymphocyte, monocyte, and granulocyte, CD4, natural killer, and B cells).^23,24^ A PCA were performed to quantify latent structures or batch effects in the data. We use the array annotations provided by Illumina to map probes to their corresponding genes.

### Statistical analysis

The EWAS between methylation level at each CpG-site (dependent variable, continuous) and each SDOH factor (independent variable, categorical) were evaluated using multiple linear regression with covariate adjustments including sex, attained age, race, cancer treatment exposures, lymphocyte subtype proportions, significant genetic principal components and methylation principal components (model 1). Health behaviors (BMI and smoking) were additionally adjusted to evaluate the contribution of each SDOH factor on methylation level independent of health behaviors (model 2). However, because health behaviors are intermediate variables and potentially on the causal pathway from SDOH factors to DNAm, we would avoid the over adjustment^25^ and treated epigenome-wide significant results from model 1 as the primary results with a post filtering step with the criteria of P<0.05 in model 2 to derive more credible SDOH-associated CpGs considering the effect did not completely disappear in model 2. We used R package CpGassoc^26^ for the linear regression analysis and used P<9×10^−8^ as the epigenome-wide significance threshold.^27^ EWAS results were visualized with circos plot (Circos v.0.69).^28^

Multivariable logistic regression models were used to assess the associations between the adjusted M-value for each SDOH-associated CpG (by the same set of covariates specified in aforementioned EWAS model) and each of pulmonary condition (binary outcome) with covariates including sex, age, BMI, smoking, and radiation to the chest (a treatment exposure significantly associated with pulmonary conditions). Only CpG-CHC associations with the significant false discovery rate (FDR) adjusted p-value (*P*_FDR_< 0.05) were regarded as statistical significance and were advanced as candidate CpGs for the downstream mediation analysis.

In the mediation analysis, each of the candidate CpGs was considered as a hypothesized mediating variable, each SDOH factor as the exposure, status of each pulmonary condition (binary) as the outcome, and adjusted for sex, age, BMI, smoking, and radiation to the chest. Individually significant mediator CpGs will be checked for pair-wise correlations. All independent significant mediator CpGs (squared Pearson correlation coefficient r^2^<0.05) were combined into a methylation risk score for a final mediation analysis. Mediation analysis was carried out using Mediation R package.^29^ All statistical analyses were performed by using R.3.6.3^30^ or SAS 9.4 (SAS Institute Inc., Cary, NC, USA) and all statistical tests were two-sided.

## Discussion

To the best of our knowledge, this is the first social epigenomic study conducted among survivors of childhood cancer. We found 130 SDOH-CpG associations for educational attainment, personal income, or ADI at epigenome-wide significance level. However, there were an order of magnitude fewer number of CpGs than those associated with cancer treatment exposures,^15^ suggesting that the SDOH factors may have smaller effect sizes compared to intensive cancer treatment modalities. Among all three SDOH factors that were examined, educational attainment seems to be the most informative factor with the largest number of significant CpG associations. Most of the CpGs were mapped to genes that were known to have DNAm associated with smoking exposure, suggesting that social and behavioral exposures to lower educational attainment, lower income and higher SES disadvantageous neighborhood resemble the effect of tobacco use. More interestingly, there were also some robust findings not related to smoking exposures. For example, cg06359375 was uniquely associated with educational attainment (but neither personal income nor area deprivation index) at epigenome-wide significant level with no attenuation for the effect size after adjusting for BMI and smoking. This CpG is mapped to *HPS4* gene, which was previously reported to be associated with cognitive function.^31^

Although biological pathways from which socio-economic vulnerability influences health outcomes in survivors of childhood cancer have not been determined, according to a biological embedding of childhood adversity model, childhood stress can get programmed into macrophage through epigenetic markings, post-translational modifications, and tissue remodeling. This could cause the immune cells to mount excessive inflammatory to microbial challenges and be insensitivity to inhibitory hormonal signals, resulting in a chronic inflammatory state in the body.^32^ Moreover, several studies have supported that SES was biologically embedded by showing that low SES across the life course has been associated with a blunted pattern of diurnal cortisol production,^33^ higher level of allostatic load,^33^ increased inflammatory activity,^34-36^ and higher pathogen burden.^37,38^

EWAS findings suggest potential implications of SDOH-associated CpGs in risk of three common pulmonary diseases that were clinically-assessed during SJLIFE follow-up visits. We found AA survivors had significantly higher incidence of pulmonary diffusion deficit or restrictive pulmonary deficit than EA survivors. In contrast, there is no racial disparity for incidence of obstructive pulmonary deficit. Considering AA survivors had worse socio-economic exposures than EA based on all three key factors (i.e., lower educational attainment, lower personal income, and higher national rank of ADI), we continued to examine whether SDOH can account for, to some extent, the observed higher incidence of pulmonary diseases. We observed two distinct patterns; the racial disparity in pulmonary diffusion deficits disappeared after adjusting for SDOH factors suggesting significant contribution of SDOH factors to racial disparity in pulmonary diffusion deficits, whereas the racial disparity in restrictive pulmonary deficits remained significant with little change.

To illuminate potential biological mechanism for the SDOH effect on pulmonary diffusion deficit, we evaluated the association between SDOH-associated CpGs and incidence of pulmonary diffusion deficit and further performed mediation analysis for a subset of CpGs (mediator) that are associated with both SDOH (exposure) and pulmonary diffusion deficit (outcome). There were three CpG collectively contributed nearly 50% mediation effect of educational attainment, and a single mediator CpG had partial mediation of personal income and ADI on pulmonary diffusion deficit. In contrast, we did not find any of these SDOH associated CpGs were associated with either obstructive pulmonary deficit or restrictive pulmonary deficit.

Our study has the following limitations; First, the sample size of the AA population was small for identification of race specific epigenetic association with SDOH factors, but 20% (26/130) epigenome-wide significant CpGs among EA survivors were validated using AA survivors (*P*<0.05). Second, the analysis was based on cross-sectional study, so there was no clearly defined temporal association to establish the causality. Ideally, DNAm measured at two time points for the same set of survivors with several years apart would allow us to more rigorously analyze the effect of baseline SDOH factors (or change of SDOH) on the changes of DNAm between the two consecutive time points and also evaluate the temporal association between DNAm measured at the earlier time point and incidence CHC occurred at later time point. Third, we attempted to take advantage of the existing whole-genome sequencing data to search for methylation quantitative trait loci (meQTL) but we did not find any strong meQTL for the top genomic region on chromosome 3 that can be used in mendelian randomization^39^ for causal inference.

In summary, the first social epigenomic study in childhood cancer survivors identified striking DNAm signatures associated with multiple SDOH factors, with many epigenome-wide significant CpG sites that mimic the effect of smoking exposure. By taking a social epigenomic approach, we also demonstrated an exemplified racial health disparity in pulmonary diffusion deficit between AA and EA survivors, and illuminated DNAm as potential mediators for SDOH factors which contributed to the disparity.

## Data Availability

The St. Jude Lifetime Cohort Study data are accessible through the St. Jude Cloud (https://stjude.cloud).

