## Supplementary Tables for "Epigenome-wide association studies of three social determinants of health and implications for lung functions among survivors of childhood cancer"

| **Supplementary Table 1. Association of DNA methylation CpG sites with social determinants of health factors in survivors of European ancestry (*P*<9×10^-8^)** | | | | | | | | | | |
| --- | --- | --- | --- | --- | --- | --- | --- | --- | --- | --- |
| Social-economic factor | CpG label | Chr | Position | HGNC gene | Model 1 | | | Model 2 (Smoking/BMI adjusted model) | | |
|  |  |  |  |  | Effect size | (SE) | *P* | Effect size | (SE) | *P* |
| Educational attainment | cg19859270 | chr3 | 98,532,449 | *CPOX;GPR15* | 0.109 | (0.008) | 4.5E-37 | 0.037 | (0.007) | 2.4E-07 |
| Educational attainment | cg05575921 | chr5 | 373,262 | *AHRR* | 0.136 | (0.010) | 8.1E-37 | 0.040 | (0.008) | 1.5E-06 |
| **Educational attainment** | **cg04180924** | **chr3** | **98,553,219** | ***CPOX*** | **0.100** | **(0.008)** | **1.9E-35** | **0.039** | **(0.007)** | **2.0E-08** |
| Educational attainment | cg02978227 | chr3 | 98,573,182 | *CPOX* | 0.080 | (0.007) | 3.5E-32 | 0.025 | (0.006) | 1.6E-05 |
| Educational attainment | cg05659611 | chr3 | 98,521,779 | *CLDND1;CPOX;RP11-227H4.5* | 0.065 | (0.005) | 5.0E-31 | 0.026 | (0.005) | 5.4E-07 |
| Educational attainment | cg02657160 | chr3 | 98,592,218 | *CPOX* | 0.055 | (0.005) | 8.9E-29 | 0.020 | (0.004) | 9.4E-06 |
| Educational attainment | cg08064403 | chr3 | 98,521,413 | *CLDND1;CPOX;RP11-227H4.5* | 0.103 | (0.009) | 3.4E-28 | 0.033 | (0.008) | 6.8E-05 |
| Educational attainment | cg26768182 | chr9 | 131,397,291 | *PRRC2B* | 0.061 | (0.006) | 4.6E-26 | 0.027 | (0.005) | 8.0E-07 |
| Educational attainment | cg00010201 | chr3 | 98,568,261 | *CPOX* | 0.081 | (0.008) | 1.4E-25 | 0.029 | (0.007) | 6.9E-05 |
| **Educational attainment** | **cg04885881** | **chr1** | **11,063,060** | ***-*** | **0.042** | **(0.004)** | **3.2E-24** | **0.026** | **(0.004)** | **1.3E-09** |
| Educational attainment | cg00385142 | chr3 | 98,517,073 | *CLDND1* | 0.039 | (0.004) | 1.8E-20 | 0.019 | (0.004) | 7.6E-06 |
| Educational attainment | cg21566642 | chr2 | 232,419,950 | *ECEL1P1* | 0.035 | (0.004) | 2.3E-20 | 0.015 | (0.004) | 8.4E-05 |
| Educational attainment | cg13025388 | chr11 | 118,228,036 | *MPZL3* | 0.051 | (0.006) | 1.7E-17 | 0.024 | (0.006) | 9.2E-05 |
| Educational attainment | cg06362176 | chr9 | 124,286,348 | *NEK6* | 0.027 | (0.003) | 7.9E-17 | 0.015 | (0.003) | 1.3E-05 |
| Educational attainment | cg03028088 | chr10 | 62,083,461 | *ARID5B* | 0.024 | (0.003) | 9.8E-16 | 0.013 | (0.003) | 2.5E-05 |
| Educational attainment | cg26878655 | chr17 | 40,325,624 | *RARA* | 0.030 | (0.004) | 2.7E-15 | 0.017 | (0.004) | 1.5E-05 |
| Educational attainment | cg12329529 | chr11 | 61,055,353 | *RP11-881M11.8* | 0.031 | (0.004) | 6.2E-14 | 0.017 | (0.004) | 8.4E-05 |
| Educational attainment | cg06807063 | chr16 | 3,596,052 | *SLX4* | 0.027 | (0.004) | 5.9E-13 | 0.020 | (0.004) | 5.3E-07 |
| Educational attainment | cg00809772 | chr11 | 6,746,852 | *GVINP1;GVINP2* | -0.029 | (0.004) | 1.3E-12 | -0.023 | (0.004) | 2.2E-07 |
| Educational attainment | cg15349661 | chr10 | 4,127,748 | *-* | 0.030 | (0.004) | 2.1E-12 | 0.018 | (0.005) | 7.5E-05 |
| Educational attainment | cg07162861 | chr15 | 90,872,777 | *FURIN* | 0.030 | (0.004) | 5.6E-12 | 0.023 | (0.005) | 9.8E-07 |
| Educational attainment | cg23662846 | chr15 | 90,872,775 | *FURIN* | 0.033 | (0.005) | 5.7E-12 | 0.024 | (0.005) | 2.8E-06 |
| Educational attainment | cg07178945 | chr12 | 4,379,633 | *FGF23* | -0.029 | (0.004) | 2.4E-11 | -0.018 | (0.005) | 9.9E-05 |
| Educational attainment | cg18896032 | chr11 | 57,330,243 | *SSRP1* | 0.030 | (0.004) | 3.0E-11 | 0.021 | (0.005) | 1.0E-05 |
| Educational attainment | cg15241876 | chr22 | 36,861,551 | *NCF4;NCF4-AS1* | 0.024 | (0.004) | 4.9E-11 | 0.016 | (0.004) | 5.1E-05 |
| Educational attainment | cg04357217 | chr12 | 12,673,617 | *GPR19;RP11-180M15.3* | 0.034 | (0.005) | 5.2E-11 | 0.022 | (0.006) | 7.2E-05 |
| Educational attainment | cg00526275 | chr12 | 118,373,745 | *TAOK3* | 0.032 | (0.005) | 5.7E-11 | 0.022 | (0.005) | 2.3E-05 |
| Educational attainment | cg26451984 | chr2 | 239,146,470 | *HDAC4* | 0.020 | (0.003) | 6.5E-11 | 0.013 | (0.003) | 1.1E-04 |
| Educational attainment | cg16509061 | chr5 | 1,269,193 | *TERT* | 0.021 | (0.003) | 1.0E-10 | 0.016 | (0.004) | 8.2E-06 |
| Educational attainment | cg17708080 | chr10 | 110,426,276 | *-* | 0.018 | (0.003) | 1.1E-10 | 0.015 | (0.003) | 9.0E-07 |
| Educational attainment | cg13518852 | chr1 | 212,718,663 | *-* | -0.034 | (0.005) | 1.5E-10 | -0.022 | (0.006) | 1.2E-04 |
| Educational attainment | cg12722937 | chr15 | 90,872,816 | *FURIN* | 0.017 | (0.003) | 1.7E-10 | 0.013 | (0.003) | 1.6E-05 |
| Educational attainment | cg11813497 | chr10 | 14,330,879 | *FRMD4A* | -0.048 | (0.007) | 1.7E-10 | -0.039 | (0.008) | 1.1E-06 |
| Educational attainment | cg05389935 | chr1 | 26,284,265 | *UBXN11* | 0.019 | (0.003) | 2.6E-10 | 0.013 | (0.003) | 3.5E-05 |
| Educational attainment | cg17955636 | chr11 | 61,055,669 | *RP11-881M11.8* | 0.020 | (0.003) | 2.6E-10 | 0.013 | (0.003) | 1.2E-04 |
| Educational attainment | cg25713841 | chr22 | 25,702,831 | *GRK3* | 0.028 | (0.004) | 2.8E-10 | 0.018 | (0.005) | 1.2E-04 |
| Educational attainment | cg03660377 | chr16 | 318,047 | *AXIN1* | 0.024 | (0.004) | 3.0E-10 | 0.016 | (0.004) | 1.2E-04 |
| Educational attainment | cg04791659 | chr15 | 90,872,581 | *FURIN* | 0.027 | (0.004) | 3.2E-10 | 0.019 | (0.005) | 7.7E-05 |
| Educational attainment | cg11935248 | chr6 | 4,942,248 | *CDYL* | 0.021 | (0.003) | 3.4E-10 | 0.014 | (0.004) | 6.6E-05 |
| Educational attainment | cg13345299 | chr22 | 26,480,108 | *HPS4* | 0.020 | (0.003) | 4.9E-10 | 0.018 | (0.003) | 3.7E-07 |
| Educational attainment | cg27596226 | chr16 | 3,595,840 | *SLX4* | 0.017 | (0.003) | 5.3E-10 | 0.014 | (0.003) | 1.3E-06 |
| Educational attainment | cg20451986 | chr11 | 134,058,406 | *-* | -0.026 | (0.004) | 5.6E-10 | -0.020 | (0.005) | 7.3E-06 |
| Educational attainment | cg00207731 | chr15 | 90,872,887 | *FURIN* | 0.024 | (0.004) | 6.4E-10 | 0.016 | (0.004) | 1.5E-04 |
| Educational attainment | cg25252598 | chr12 | 6,532,435 | *GAPDH;RP5-940J5.3* | 0.021 | (0.003) | 7.8E-10 | 0.014 | (0.004) | 1.4E-04 |
| Educational attainment | cg13410000 | chr15 | 90,814,290 | *BLM* | 0.023 | (0.004) | 8.3E-10 | 0.018 | (0.004) | 7.7E-06 |
| Educational attainment | cg12336290 | chr10 | 70,602,917 | *PRF1* | -0.023 | (0.004) | 1.0E-09 | -0.016 | (0.004) | 1.1E-04 |
| Educational attainment | cg15324996 | chr3 | 187,975,557 | *RP11-132N15.3* | 0.020 | (0.003) | 1.2E-09 | 0.014 | (0.004) | 6.1E-05 |
| Educational attainment | cg02532700 | chr22 | 36,861,361 | *NCF4;NCF4-AS1* | 0.030 | (0.005) | 1.3E-09 | 0.022 | (0.005) | 4.1E-05 |
| Educational attainment | cg20208538 | chr3 | 20,054,853 | *KAT2B* | 0.029 | (0.005) | 1.6E-09 | 0.024 | (0.005) | 1.0E-05 |
| Educational attainment | cg02223801 | chr7 | 157,574,240 | *PTPRN2* | 0.026 | (0.004) | 1.6E-09 | 0.018 | (0.005) | 9.9E-05 |
| Educational attainment | cg07013698 | chr17 | 82,216,903 | *RP13-516M14.2* | 0.012 | (0.002) | 1.8E-09 | 0.008 | (0.002) | 1.2E-04 |
| Educational attainment | cg27161340 | chr16 | 87,459,065 | *ZCCHC14* | 0.019 | (0.003) | 1.8E-09 | 0.015 | (0.003) | 9.2E-06 |
| Educational attainment | cg23670794 | chr3 | 111,594,217 | *CD96;ZBED2* | 0.026 | (0.004) | 2.2E-09 | 0.018 | (0.005) | 1.1E-04 |
| Educational attainment | cg07742906 | chr1 | 11,883,373 | *-* | -0.018 | (0.003) | 2.4E-09 | -0.013 | (0.003) | 6.7E-05 |
| Educational attainment | cg18274515 | chr15 | 90,872,301 | *FURIN* | 0.020 | (0.003) | 2.6E-09 | 0.016 | (0.004) | 3.0E-05 |
| Educational attainment | cg13310423 | chr2 | 96,147,050 | *AC012307.2* | 0.015 | (0.003) | 3.3E-09 | 0.012 | (0.003) | 3.3E-05 |
| Educational attainment | cg27251814 | chr22 | 26,479,891 | *HPS4* | 0.026 | (0.004) | 3.4E-09 | 0.023 | (0.005) | 2.4E-06 |
| Educational attainment | cg24931191 | chr3 | 36,207,366 | *-* | -0.037 | (0.006) | 4.0E-09 | -0.030 | (0.007) | 8.6E-06 |
| Educational attainment | cg10698959 | chr15 | 90,872,829 | *FURIN* | 0.016 | (0.003) | 4.1E-09 | 0.013 | (0.003) | 3.2E-05 |
| Educational attainment | cg08214029 | chr17 | 36,064,117 | *CCL18* | 0.015 | (0.003) | 4.2E-09 | 0.011 | (0.003) | 1.1E-04 |
| **Educational attainment** | **cg06359375** | **chr22** | **26,479,837** | ***HPS4*** | **0.023** | **(0.004)** | **4.3E-09** | **0.023** | **(0.004)** | **2.0E-08** |
| Educational attainment | cg07958818 | chr1 | 19,440,699 | *CAPZB* | 0.020 | (0.003) | 4.4E-09 | 0.015 | (0.004) | 6.7E-05 |
| Educational attainment | cg20185195 | chr1 | 38,929,700 | *RHBDL2* | 0.022 | (0.004) | 4.6E-09 | 0.016 | (0.004) | 6.7E-05 |
| Educational attainment | cg26669717 | chr7 | 601,390 | *PRKAR1B* | 0.015 | (0.003) | 4.9E-09 | 0.011 | (0.003) | 1.4E-04 |
| Educational attainment | cg21431832 | chr22 | 26,479,685 | *HPS4* | 0.025 | (0.004) | 6.0E-09 | 0.023 | (0.005) | 1.5E-06 |
| Educational attainment | cg27470486 | chr17 | 41,917,434 | *ACLY* | 0.015 | (0.003) | 6.6E-09 | 0.011 | (0.003) | 7.5E-05 |
| Educational attainment | cg23059461 | chr10 | 70,602,937 | *PRF1* | -0.023 | (0.004) | 8.4E-09 | -0.016 | (0.004) | 1.2E-04 |
| Educational attainment | cg12624850 | chr19 | 16,120,202 | *RAB8A* | 0.017 | (0.003) | 9.2E-09 | 0.013 | (0.003) | 5.0E-05 |
| Educational attainment | cg27468704 | chr22 | 26,479,856 | *HPS4* | 0.028 | (0.005) | 1.1E-08 | 0.027 | (0.005) | 2.7E-07 |
| Educational attainment | cg15976297 | chr22 | 45,996,348 | *-* | 0.013 | (0.002) | 1.3E-08 | 0.009 | (0.002) | 9.9E-05 |
| Educational attainment | cg01139665 | chr19 | 4,841,343 | *PLIN3* | 0.019 | (0.003) | 1.3E-08 | 0.017 | (0.004) | 3.3E-06 |
| Educational attainment | cg21159885 | chr1 | 92,978,146 | *-* | 0.026 | (0.005) | 1.4E-08 | 0.020 | (0.005) | 1.0E-04 |
| Educational attainment | cg18282388 | chr11 | 129,846,574 | *TMEM45B* | 0.025 | (0.004) | 2.1E-08 | 0.019 | (0.005) | 3.8E-05 |
| Educational attainment | cg06483559 | chr11 | 123,754,205 | *OR6X1* | 0.014 | (0.003) | 2.2E-08 | 0.011 | (0.003) | 4.7E-05 |
| Educational attainment | cg27614723 | chr15 | 91,856,666 | *SLCO3A1* | 0.015 | (0.003) | 3.1E-08 | 0.011 | (0.003) | 1.0E-04 |
| Educational attainment | cg25464840 | chr10 | 14,330,910 | *FRMD4A* | -0.032 | (0.006) | 3.1E-08 | -0.025 | (0.006) | 8.2E-05 |
| Educational attainment | cg15088991 | chr1 | 26,791,730 | *PIGV* | 0.019 | (0.003) | 3.2E-08 | 0.015 | (0.004) | 6.5E-05 |
| Educational attainment | cg22543377 | chr10 | 113,170,419 | *-* | 0.024 | (0.004) | 3.6E-08 | 0.021 | (0.005) | 1.1E-05 |
| Educational attainment | cg22335340 | chr12 | 6,945,989 | *C12orf57;PTPN6;U47924.31* | 0.015 | (0.003) | 4.0E-08 | 0.011 | (0.003) | 1.3E-04 |
| Educational attainment | cg07915117 | chr1 | 31,955,529 | *-* | 0.023 | (0.004) | 5.6E-08 | 0.018 | (0.005) | 8.4E-05 |
| Educational attainment | cg03121287 | chr5 | 150,155,823 | *PDGFRB* | -0.015 | (0.003) | 5.7E-08 | -0.014 | (0.003) | 5.9E-06 |
| Educational attainment | cg11205006 | chr22 | 26,479,532 | *HPS4* | 0.026 | (0.005) | 5.9E-08 | 0.024 | (0.005) | 6.4E-06 |
| Educational attainment | cg09006159 | chr2 | 97,713,347 | *ZAP70* | -0.018 | (0.003) | 6.2E-08 | -0.014 | (0.004) | 7.3E-05 |
| Educational attainment | cg13658022 | chr2 | 241,376,960 | *FARP2* | 0.018 | (0.003) | 6.7E-08 | 0.014 | (0.004) | 9.2E-05 |
| Educational attainment | cg06648759 | chr13 | 40,318,613 | *-* | -0.019 | (0.003) | 7.0E-08 | -0.016 | (0.004) | 2.1E-05 |
| Educational attainment | cg15507334 | chr10 | 14,330,913 | *FRMD4A* | -0.029 | (0.005) | 7.2E-08 | -0.023 | (0.006) | 1.3E-04 |
| Educational attainment | cg06780383 | chr14 | 91,524,119 | *-* | -0.015 | (0.003) | 7.4E-08 | -0.012 | (0.003) | 5.0E-05 |
| Educational attainment | cg03198066 | chr2 | 158,454,555 | *CCDC148* | -0.011 | (0.002) | 8.0E-08 | -0.009 | (0.002) | 3.4E-05 |
| **Personal income** | **cg04180924** | **chr3** | **98,553,219** | ***CPOX*** | **0.069** | **(0.007)** | **3.8E-21** | **0.037** | **(0.006)** | **5.4E-09** |
| Personal income | cg05575921 | chr5 | 373,262 | *AHRR* | 0.087 | (0.010) | 1.3E-18 | 0.036 | (0.008) | 1.4E-06 |
| Personal income | cg19859270 | chr3 | 98,532,449 | *CPOX;GPR15* | 0.067 | (0.008) | 8.7E-18 | 0.029 | (0.006) | 7.6E-06 |
| Personal income | cg02657160 | chr3 | 98,592,218 | *CPOX* | 0.037 | (0.004) | 2.0E-16 | 0.018 | (0.004) | 1.2E-05 |
| Personal income | cg08064403 | chr3 | 98,521,413 | *CLDND1;CPOX;RP11-227H4.5* | 0.070 | (0.008) | 2.6E-16 | 0.034 | (0.008) | 5.9E-06 |
| Personal income | cg02978227 | chr3 | 98,573,182 | *CPOX* | 0.048 | (0.006) | 8.1E-15 | 0.020 | (0.005) | 1.8E-04 |
| Personal income | cg18754985 | chr3 | 98,518,905 | *CLDND1* | 0.037 | (0.005) | 4.6E-14 | 0.019 | (0.005) | 5.5E-05 |
| Personal income | cg04885881 | chr1 | 11,063,060 | *-* | 0.028 | (0.004) | 1.1E-13 | 0.020 | (0.004) | 4.6E-07 |
| Personal income | cg05659611 | chr3 | 98,521,779 | *CLDND1;CPOX;RP11-227H4.5* | 0.038 | (0.005) | 1.9E-13 | 0.019 | (0.005) | 7.4E-05 |
| Personal income | cg00010201 | chr3 | 98,568,261 | *CPOX* | 0.049 | (0.007) | 2.1E-12 | 0.022 | (0.007) | 8.8E-04 |
| Personal income | cg00385142 | chr3 | 98,517,073 | *CLDND1* | 0.025 | (0.004) | 1.0E-10 | 0.014 | (0.004) | 1.6E-04 |
| Personal income | cg01940273 | chr2 | 232,420,223 | *ECEL1P1* | 0.030 | (0.005) | 4.0E-10 | 0.016 | (0.005) | 6.1E-04 |
| Personal income | cg06035956 | chr5 | 378,983 | *AHRR* | 0.027 | (0.004) | 8.1E-10 | 0.015 | (0.004) | 5.5E-04 |
| Personal income | cg27521648 | chr3 | 98,574,171 | *CPOX* | 0.026 | (0.004) | 9.8E-10 | 0.015 | (0.004) | 6.1E-04 |
| Personal income | cg06805218 | chr5 | 14,676,513 | *OTULIN* | 0.020 | (0.003) | 1.4E-09 | 0.014 | (0.003) | 6.0E-05 |
| Personal income | cg23608915 | chr2 | 101,717,332 | *MAP4K4* | -0.022 | (0.004) | 9.1E-09 | -0.019 | (0.004) | 4.0E-06 |
| Personal income | cg09422787 | chr14 | 73,741,152 | *ELMSAN1* | 0.011 | (0.002) | 1.1E-08 | 0.009 | (0.002) | 1.9E-05 |
| Personal income | cg11063902 | chr3 | 100,074,148 | *CMSS1;FILIP1L* | 0.021 | (0.004) | 1.6E-08 | 0.014 | (0.004) | 1.9E-04 |
| Personal income | cg06235438 | chr16 | 30,474,644 | *ITGAL* | 0.019 | (0.003) | 2.8E-08 | 0.011 | (0.003) | 1.1E-03 |
| Personal income | cg20650823 | chr14 | 102,479,844 | *TECPR2* | -0.020 | (0.004) | 3.7E-08 | -0.017 | (0.004) | 2.0E-05 |
| Personal income | cg16529797 | chr8 | 133,133,207 | *TG* | 0.017 | (0.003) | 4.7E-08 | 0.014 | (0.003) | 3.0E-05 |
| Personal income | cg17573365 | chr11 | 66,100,688 | *PACS1* | -0.019 | (0.004) | 5.7E-08 | -0.017 | (0.004) | 1.6E-05 |
| Personal income | cg13751113 | chr11 | 118,214,498 | *JAML* | 0.019 | (0.003) | 5.7E-08 | 0.013 | (0.004) | 3.5E-04 |
| Area deprivation index | cg05659611 | chr3 | 98,521,779 | *CLDND1;CPOX;RP11-227H4.5* | -0.002 | (0.000) | 1.3E-17 | -0.001 | (0.000) | 1.0E-06 |
| Area deprivation index | cg02657160 | chr3 | 98,592,218 | *CPOX* | -0.002 | (0.000) | 7.9E-15 | -0.001 | (0.000) | 1.6E-05 |
| Area deprivation index | cg19859270 | chr3 | 98,532,449 | *CPOX;GPR15* | -0.003 | (0.000) | 4.4E-14 | -0.001 | (0.000) | 1.0E-03 |
| Area deprivation index | cg00385142 | chr3 | 98,517,073 | *CLDND1* | -0.002 | (0.000) | 7.1E-14 | -0.001 | (0.000) | 2.3E-06 |
| Area deprivation index | cg05575921 | chr5 | 373,262 | *AHRR* | -0.004 | (0.001) | 2.5E-13 | -0.001 | (0.000) | 4.0E-04 |
| Area deprivation index | cg08064403 | chr3 | 98,521,413 | *CLDND1;CPOX;RP11-227H4.5* | -0.003 | (0.000) | 1.3E-12 | -0.001 | (0.000) | 6.6E-04 |
| Area deprivation index | cg02978227 | chr3 | 98,573,182 | *CPOX* | -0.002 | (0.000) | 1.8E-12 | -0.001 | (0.000) | 2.0E-03 |
| Area deprivation index | cg04180924 | chr3 | 98,553,219 | *CPOX* | -0.003 | (0.000) | 4.4E-12 | -0.001 | (0.000) | 1.5E-03 |
| Area deprivation index | cg12127149 | chr14 | 102,020,682 | *DYNC1H1* | -0.001 | (0.000) | 2.0E-09 | -0.001 | (0.000) | 7.2E-06 |
| Area deprivation index | cg12803068 | chr7 | 44,963,319 | *MYO1G* | 0.003 | (0.001) | 5.3E-09 | 0.002 | (0.001) | 3.6E-04 |
| Area deprivation index | cg01899089 | chr5 | 369,853 | *AHRR* | -0.001 | (0.000) | 5.4E-09 | 0.000 | (0.000) | 1.6E-03 |
| Area deprivation index | cg05221370 | chr7 | 111,098,779 | *IMMP2L;LRRN3* | -0.002 | (0.000) | 3.3E-08 | -0.001 | (0.000) | 7.6E-04 |
| Area deprivation index | cg08840017 | chr2 | 230,944,894 | *GPR55* | -0.001 | (0.000) | 4.0E-08 | -0.001 | (0.000) | 1.4E-03 |
| Area deprivation index | cg04180046 | chr7 | 44,963,136 | *MYO1G* | 0.002 | (0.000) | 5.3E-08 | 0.001 | (0.000) | 2.8E-04 |
| Area deprivation index | cg23662846 | chr15 | 90,872,775 | *FURIN* | -0.001 | (0.000) | 5.8E-08 | -0.001 | (0.000) | 1.4E-04 |
| Area deprivation index | cg05445320 | chr20 | 47,356,991 | *ZMYND8* | -0.001 | (0.000) | 6.1E-08 | -0.001 | (0.000) | 4.3E-05 |
| Area deprivation index | cg01904393 | chr12 | 51,421,298 | *SLC4A8* | -0.001 | (0.000) | 6.2E-08 | -0.001 | (0.000) | 2.1E-05 |
| Area deprivation index | cg26577523 | chr2 | 70,043,693 | *PCBP1-AS1* | -0.001 | (0.000) | 6.4E-08 | 0.000 | (0.000) | 2.1E-03 |
| Area deprivation index | cg17585440 | chr19 | 2,052,181 | *MKNK2* | -0.001 | (0.000) | 8.5E-08 | 0.000 | (0.000) | 5.7E-05 |
| Abbreviations: Chr (chromosome), HGNC (The HUGO genome nomenclature committee), SE (standard error) | | | | | | | | | | |

| **Supplementary Table 2. Association of DNA methylation CpG sites with social determinants of health factors in survivors of African ancestry** | | | | | | | |
| --- | --- | --- | --- | --- | --- | --- | --- |
| Social determinants of health | CpG label | Chr | Position | HGNC gene | Effect size | (SE) | *P* |
| Educational attainment | cg19859270 | chr3 | 98,532,449 | *CPOX;GPR15* | 0.0827 | (0.0241) | 0.0007 |
| Educational attainment | cg05575921 | chr5 | 373,262 | *AHRR* | 0.1059 | (0.0316) | 0.0009 |
| Educational attainment | cg04180924 | chr3 | 98,553,219 | *CPOX* | 0.0617 | (0.0215) | 0.0045 |
| Educational attainment | cg02978227 | chr3 | 98,573,182 | *CPOX* | 0.0715 | (0.0204) | 0.0005 |
| Educational attainment | cg05659611 | chr3 | 98,521,779 | *CLDND1;CPOX;RP11-227H4.5* | 0.0507 | (0.0146) | 0.0006 |
| Educational attainment | cg02657160 | chr3 | 98,592,218 | *CPOX* | 0.0332 | (0.0129) | 0.0107 |
| Educational attainment | cg08064403 | chr3 | 98,521,413 | *CLDND1;CPOX;RP11-227H4.5* | 0.0575 | (0.0254) | 0.0245 |
| Educational attainment | cg26768182 | chr9 | 131,397,291 | *PRRC2B* | 0.0347 | (0.0161) | 0.0322 |
| Educational attainment | cg00010201 | chr3 | 98,568,261 | *CPOX* | 0.0381 | (0.0272) | 0.1619 |
| Educational attainment | cg04885881 | chr1 | 11,063,060 | *-* | 0.0327 | (0.0119) | 0.0067 |
| Educational attainment | cg00385142 | chr3 | 98,517,073 | *CLDND1* | 0.0193 | (0.0106) | 0.0702 |
| Educational attainment | cg21566642 | chr2 | 232,419,950 | *ECEL1P1* | 0.0336 | (0.0133) | 0.0122 |
| Educational attainment | cg13025388 | chr11 | 118,228,036 | *MPZL3* | 0.0112 | (0.0169) | 0.5104 |
| Educational attainment | cg06362176 | chr9 | 124,286,348 | *NEK6* | 0.0156 | (0.0098) | 0.1123 |
| Educational attainment | cg03028088 | chr10 | 62,083,461 | *ARID5B* | 0.0111 | (0.0092) | 0.2295 |
| Educational attainment | cg26878655 | chr17 | 40,325,624 | *RARA* | 0.0138 | (0.0106) | 0.1952 |
| Educational attainment | cg12329529 | chr11 | 61,055,353 | *RP11-881M11.8* | -0.0132 | (0.0135) | 0.3284 |
| Educational attainment | cg06807063 | chr16 | 3,596,052 | *SLX4* | 0.0478 | (0.0442) | 0.2802 |
| Educational attainment | cg00809772 | chr11 | 6,746,852 | *GVINP1;GVINP2* | -0.0010 | (0.0152) | 0.9456 |
| Educational attainment | cg15349661 | chr10 | 4,127,748 | *-* | 0.0308 | (0.0138) | 0.0268 |
| Educational attainment | cg07162861 | chr15 | 90,872,777 | *FURIN* | 0.0225 | (0.0118) | 0.0588 |
| Educational attainment | cg23662846 | chr15 | 90,872,775 | *FURIN* | 0.0153 | (0.0138) | 0.2687 |
| Educational attainment | cg07178945 | chr12 | 4,379,633 | *FGF23* | -0.0197 | (0.0119) | 0.1007 |
| Educational attainment | cg18896032 | chr11 | 57,330,243 | *SSRP1* | 0.0099 | (0.0120) | 0.4107 |
| Educational attainment | cg15241876 | chr22 | 36,861,551 | *NCF4;NCF4-AS1* | 0.0128 | (0.0100) | 0.2009 |
| Educational attainment | cg04357217 | chr12 | 12,673,617 | *GPR19;RP11-180M15.3* | -0.0036 | (0.0158) | 0.8196 |
| Educational attainment | cg00526275 | chr12 | 118,373,745 | *TAOK3* | 0.0287 | (0.0190) | 0.1325 |
| Educational attainment | cg26451984 | chr2 | 239,146,470 | *HDAC4* | 0.0162 | (0.0094) | 0.0843 |
| Educational attainment | cg16509061 | chr5 | 1,269,193 | *TERT* | -0.0019 | (0.0099) | 0.8454 |
| Educational attainment | cg17708080 | chr10 | 110,426,276 | *-* | -0.0055 | (0.0080) | 0.4904 |
| Educational attainment | cg13518852 | chr1 | 212,718,663 | *-* | 0.0207 | (0.0161) | 0.2014 |
| Educational attainment | cg12722937 | chr15 | 90,872,816 | *FURIN* | 0.0125 | (0.0072) | 0.0830 |
| Educational attainment | cg11813497 | chr10 | 14,330,879 | *FRMD4A* | -0.0218 | (0.0216) | 0.3143 |
| Educational attainment | cg05389935 | chr1 | 26,284,265 | *UBXN11* | 0.0015 | (0.0083) | 0.8581 |
| Educational attainment | cg17955636 | chr11 | 61,055,669 | *RP11-881M11.8* | -0.0140 | (0.0092) | 0.1311 |
| Educational attainment | cg25713841 | chr22 | 25,702,831 | *GRK3* | 0.0124 | (0.0129) | 0.3373 |
| Educational attainment | cg03660377 | chr16 | 318,047 | *AXIN1* | 0.0066 | (0.0119) | 0.5808 |
| Educational attainment | cg04791659 | chr15 | 90,872,581 | *FURIN* | 0.0175 | (0.0128) | 0.1731 |
| Educational attainment | cg11935248 | chr6 | 4,942,248 | *CDYL* | 0.0135 | (0.0091) | 0.1387 |
| Educational attainment | cg13345299 | chr22 | 26,480,108 | *HPS4* | 0.0126 | (0.0100) | 0.2067 |
| Educational attainment | cg27596226 | chr16 | 3,595,840 | *SLX4* | -0.0133 | (0.0085) | 0.1192 |
| Educational attainment | cg20451986 | chr11 | 134,058,406 | *-* | -0.0037 | (0.0113) | 0.7410 |
| Educational attainment | cg00207731 | chr15 | 90,872,887 | *FURIN* | 0.0136 | (0.0105) | 0.1982 |
| Educational attainment | cg25252598 | chr12 | 6,532,435 | *GAPDH;RP5-940J5.3* | -0.0025 | (0.0111) | 0.8220 |
| Educational attainment | cg13410000 | chr15 | 90,814,290 | *BLM* | 0.0154 | (0.0108) | 0.1579 |
| Educational attainment | cg12336290 | chr10 | 70,602,917 | *PRF1* | 0.0004 | (0.0119) | 0.9743 |
| Educational attainment | cg15324996 | chr3 | 187,975,557 | *RP11-132N15.3* | 0.0176 | (0.0096) | 0.0699 |
| Educational attainment | cg02532700 | chr22 | 36,861,361 | *NCF4;NCF4-AS1* | 0.0147 | (0.0145) | 0.3115 |
| Educational attainment | cg20208538 | chr3 | 20,054,853 | *KAT2B* | 0.0014 | (0.0133) | 0.9170 |
| Educational attainment | cg02223801 | chr7 | 157,574,240 | *PTPRN2* | -0.0103 | (0.0150) | 0.4912 |
| Educational attainment | cg07013698 | chr17 | 82,216,903 | *RP13-516M14.2* | 0.0112 | (0.0056) | 0.0448 |
| Educational attainment | cg27161340 | chr16 | 87,459,065 | *ZCCHC14* | -0.0070 | (0.0091) | 0.4436 |
| Educational attainment | cg23670794 | chr3 | 111,594,217 | *CD96;ZBED2* | 0.0195 | (0.0122) | 0.1118 |
| Educational attainment | cg07742906 | chr1 | 11,883,373 | *-* | -0.0246 | (0.0104) | 0.0187 |
| Educational attainment | cg18274515 | chr15 | 90,872,301 | *FURIN* | 0.0145 | (0.0088) | 0.0999 |
| Educational attainment | cg13310423 | chr2 | 96,147,050 | *AC012307.2* | 0.0071 | (0.0070) | 0.3092 |
| Educational attainment | cg27251814 | chr22 | 26,479,891 | *HPS4* | -0.0001 | (0.0119) | 0.9953 |
| Educational attainment | cg24931191 | chr3 | 36,207,366 | *-* | -0.0507 | (0.0203) | 0.0132 |
| Educational attainment | cg10698959 | chr15 | 90,872,829 | *FURIN* | 0.0046 | (0.0076) | 0.5488 |
| Educational attainment | cg08214029 | chr17 | 36,064,117 | *CCL18* | 0.0112 | (0.0077) | 0.1469 |
| Educational attainment | cg06359375 | chr22 | 26,479,837 | *HPS4* | -0.0124 | (0.0144) | 0.3903 |
| Educational attainment | cg07958818 | chr1 | 19,440,699 | *CAPZB* | 0.0129 | (0.0103) | 0.2098 |
| Educational attainment | cg20185195 | chr1 | 38,929,700 | *RHBDL2* | 0.0019 | (0.0105) | 0.8566 |
| Educational attainment | cg26669717 | chr7 | 601,390 | *PRKAR1B* | 0.0008 | (0.0074) | 0.9140 |
| Educational attainment | cg21431832 | chr22 | 26,479,685 | *HPS4* | 0.0101 | (0.0113) | 0.3728 |
| Educational attainment | cg27470486 | chr17 | 41,917,434 | *ACLY* | 0.0120 | (0.0076) | 0.1151 |
| Educational attainment | cg23059461 | chr10 | 70,602,937 | *PRF1* | 0.0021 | (0.0136) | 0.8768 |
| Educational attainment | cg12624850 | chr19 | 16,120,202 | *RAB8A* | 0.0041 | (0.0087) | 0.6371 |
| Educational attainment | cg27468704 | chr22 | 26,479,856 | *HPS4* | -0.0017 | (0.0143) | 0.9074 |
| Educational attainment | cg15976297 | chr22 | 45,996,348 | *-* | 0.0008 | (0.0075) | 0.9142 |
| Educational attainment | cg01139665 | chr19 | 4,841,343 | *PLIN3* | 0.0278 | (0.0167) | 0.0972 |
| Educational attainment | cg21159885 | chr1 | 92,978,146 | *-* | -0.0051 | (0.0132) | 0.7014 |
| Educational attainment | cg18282388 | chr11 | 129,846,574 | *TMEM45B* | -0.0042 | (0.0132) | 0.7522 |
| Educational attainment | cg06483559 | chr11 | 123,754,205 | *OR6X1* | 0.0009 | (0.0077) | 0.9056 |
| Educational attainment | cg27614723 | chr15 | 91,856,666 | *SLCO3A1* | 0.0031 | (0.0110) | 0.7804 |
| Educational attainment | cg25464840 | chr10 | 14,330,910 | *FRMD4A* | -0.0254 | (0.0167) | 0.1297 |
| Educational attainment | cg15088991 | chr1 | 26,791,730 | *PIGV* | 0.0142 | (0.0095) | 0.1362 |
| Educational attainment | cg22543377 | chr10 | 113,170,419 | *-* | 0.0207 | (0.0113) | 0.0684 |
| Educational attainment | cg22335340 | chr12 | 6,945,989 | *C12orf57;PTPN6;U47924.31* | 0.0035 | (0.0084) | 0.6795 |
| Educational attainment | cg07915117 | chr1 | 31,955,529 | *-* | 0.0090 | (0.0108) | 0.4078 |
| Educational attainment | cg03121287 | chr5 | 150,155,823 | *PDGFRB* | 0.0081 | (0.0077) | 0.2964 |
| Educational attainment | cg11205006 | chr22 | 26,479,532 | *HPS4* | 0.0054 | (0.0119) | 0.6520 |
| Educational attainment | cg09006159 | chr2 | 97,713,347 | *ZAP70* | -0.0033 | (0.0091) | 0.7143 |
| Educational attainment | cg13658022 | chr2 | 241,376,960 | *FARP2* | -0.0001 | (0.0096) | 0.9911 |
| Educational attainment | cg06648759 | chr13 | 40,318,613 | *-* | -0.0102 | (0.0159) | 0.5200 |
| Educational attainment | cg15507334 | chr10 | 14,330,913 | *FRMD4A* | -0.0308 | (0.0154) | 0.0464 |
| Educational attainment | cg06780383 | chr14 | 91,524,119 | *-* | -0.0102 | (0.0071) | 0.1508 |
| Educational attainment | cg03198066 | chr2 | 158,454,555 | *CCDC148* | 0.0011 | (0.0058) | 0.8561 |
| Personal income | cg04180924 | chr3 | 98,553,219 | *CPOX* | 0.0377 | (0.0191) | 0.0493 |
| Personal income | cg05575921 | chr5 | 373,262 | *AHRR* | 0.0822 | (0.0283) | 0.0040 |
| Personal income | cg19859270 | chr3 | 98,532,449 | *CPOX;GPR15* | 0.0466 | (0.0213) | 0.0298 |
| Personal income | cg02657160 | chr3 | 98,592,218 | *CPOX* | 0.0231 | (0.0114) | 0.0444 |
| Personal income | cg08064403 | chr3 | 98,521,413 | *CLDND1;CPOX;RP11-227H4.5* | 0.0452 | (0.0219) | 0.0397 |
| Personal income | cg02978227 | chr3 | 98,573,182 | *CPOX* | 0.0472 | (0.0177) | 0.0083 |
| Personal income | cg18754985 | chr3 | 98,518,905 | *CLDND1* | -0.0014 | (0.0125) | 0.9126 |
| Personal income | cg04885881 | chr1 | 11,063,060 | *-* | 0.0085 | (0.0108) | 0.4308 |
| Personal income | cg05659611 | chr3 | 98,521,779 | *CLDND1;CPOX;RP11-227H4.5* | 0.0302 | (0.0129) | 0.0205 |
| Personal income | cg00010201 | chr3 | 98,568,261 | *CPOX* | 0.0346 | (0.0239) | 0.1494 |
| Personal income | cg00385142 | chr3 | 98,517,073 | *CLDND1* | 0.0084 | (0.0094) | 0.3688 |
| Personal income | cg01940273 | chr2 | 232,420,223 | *ECEL1P1* | 0.0249 | (0.0141) | 0.0781 |
| Personal income | cg06035956 | chr5 | 378,983 | *AHRR* | 0.0104 | (0.0119) | 0.3833 |
| Personal income | cg27521648 | chr3 | 98,574,171 | *CPOX* | 0.0120 | (0.0103) | 0.2457 |
| Personal income | cg06805218 | chr5 | 14,676,513 | *OTULIN* | 0.0025 | (0.0092) | 0.7843 |
| Personal income | cg23608915 | chr2 | 101,717,332 | *MAP4K4* | -0.0031 | (0.0122) | 0.8019 |
| Personal income | cg09422787 | chr14 | 73,741,152 | *ELMSAN1* | -0.0002 | (0.0057) | 0.9663 |
| Personal income | cg11063902 | chr3 | 100,074,148 | *CMSS1;FILIP1L* | 0.0072 | (0.0172) | 0.6753 |
| Personal income | cg06235438 | chr16 | 30,474,644 | *ITGAL* | 0.0036 | (0.0080) | 0.6559 |
| Personal income | cg20650823 | chr14 | 102,479,844 | *TECPR2* | -0.0067 | (0.0089) | 0.4492 |
| Personal income | cg16529797 | chr8 | 133,133,207 | *TG* | 0.0073 | (0.0083) | 0.3820 |
| Personal income | cg17573365 | chr11 | 66,100,688 | *PACS1* | -0.0264 | (0.0105) | 0.0125 |
| Personal income | cg13751113 | chr11 | 118,214,498 | *JAML* | 0.0124 | (0.0110) | 0.2624 |
| Area deprivation index | cg05659611 | chr3 | 98,521,779 | *CLDND1;CPOX;RP11-227H4.5* | -0.0007 | (0.0007) | 0.2963 |
| Area deprivation index | cg02657160 | chr3 | 98,592,218 | *CPOX* | -0.0008 | (0.0006) | 0.1683 |
| Area deprivation index | cg19859270 | chr3 | 98,532,449 | *CPOX;GPR15* | -0.0024 | (0.0011) | 0.0326 |
| Area deprivation index | cg00385142 | chr3 | 98,517,073 | *CLDND1* | -0.0007 | (0.0005) | 0.1797 |
| Area deprivation index | cg05575921 | chr5 | 373,262 | *AHRR* | -0.0040 | (0.0015) | 0.0084 |
| Area deprivation index | cg08064403 | chr3 | 98,521,413 | *CLDND1;CPOX;RP11-227H4.5* | -0.0012 | (0.0011) | 0.2766 |
| Area deprivation index | cg02978227 | chr3 | 98,573,182 | *CPOX* | -0.0015 | (0.0010) | 0.1089 |
| Area deprivation index | cg04180924 | chr3 | 98,553,219 | *CPOX* | -0.0015 | (0.0010) | 0.1319 |
| Area deprivation index | cg12127149 | chr14 | 102,020,682 | *DYNC1H1* | -0.0007 | (0.0004) | 0.0872 |
| Area deprivation index | cg12803068 | chr7 | 44,963,319 | *MYO1G* | 0.0018 | (0.0013) | 0.1740 |
| Area deprivation index | cg01899089 | chr5 | 369,853 | *AHRR* | -0.0011 | (0.0004) | 0.0075 |
| Area deprivation index | cg05221370 | chr7 | 111,098,779 | *IMMP2L;LRRN3* | -0.0009 | (0.0009) | 0.2705 |
| Area deprivation index | cg08840017 | chr2 | 230,944,894 | *GPR55* | -0.0003 | (0.0007) | 0.6761 |
| Area deprivation index | cg04180046 | chr7 | 44,963,136 | *MYO1G* | 0.0003 | (0.0010) | 0.7883 |
| Area deprivation index | cg23662846 | chr15 | 90,872,775 | *FURIN* | 0.0002 | (0.0007) | 0.7520 |
| Area deprivation index | cg05445320 | chr20 | 47,356,991 | *ZMYND8* | -0.0004 | (0.0006) | 0.5255 |
| Area deprivation index | cg01904393 | chr12 | 51,421,298 | *SLC4A8* | 0.0000 | (0.0005) | 0.9886 |
| Area deprivation index | cg26577523 | chr2 | 70,043,693 | *PCBP1-AS1* | -0.0006 | (0.0005) | 0.2178 |
| Area deprivation index | cg17585440 | chr19 | 2,052,181 | *MKNK2* | -0.0001 | (0.0003) | 0.5935 |
| Abbreviations: Chr (chromosome), HGNC (The HUGO genome nomenclature committee), SE (standard error) | | | | | | | |

| **Supplementary Table 3. Multivariable analysis of each pulmonary condition risk by inclusion/exclusion of adjustments for social determinants of health in the model** | | | | |
| --- | --- | --- | --- | --- |
| Variable^*^ | Pulmonary condition | Effect size | (SE) | *P* |
| ***Basic model*** | Obstructive pulmonary deficits |  |  |  |
| (Intercept) |  | -2.987 | (0.512) | 5.3E-09 |
| Attained age, years |  | 0.008 | (0.010) | 4.2E-01 |
| Gender, female |  | -0.105 | (0.175) | 5.5E-01 |
| BMI, <18.5 |  | -0.187 | (0.477) | 6.9E-01 |
| BMI, <18.5-<25.0 |  | -0.059 | (0.211) | 7.8E-01 |
| BMI, ≥25.0 |  | -0.300 | (0.218) | 1.7E-01 |
| Past smokers |  | 0.279 | (0.226) | 2.2E-01 |
| Current smokers |  | 0.350 | (0.207) | 9.1E-02 |
| Chest RT |  | 1.184 | (0.174) | 9.7E-12 |
| Race |  | 0.235 | (0.286) | 4.1E-01 |
| ***Full model additionally adjusted for SDOH*** | Obstructive pulmonary deficits |  |  |  |
| (Intercept) |  | -2.142 | (0.761) | 4.9E-03 |
| Attained age, years |  | 0.012 | (0.012) | 3.1E-01 |
| Gender, female |  | 0.001 | (0.208) | 1.0E+00 |
| Educational attainment |  | -0.399 | (0.101) | 7.6E-05 |
| Personal income |  | 0.007 | (0.093) | 9.4E-01 |
| Area deprivation index, national rank |  | 0.002 | (0.004) | 6.7E-01 |
| BMI, <18.5 |  | -0.030 | (0.498) | 9.5E-01 |
| BMI, <18.5-<25.0 |  | 0.125 | (0.244) | 6.1E-01 |
| BMI, ≥25.0 |  | -0.353 | (0.254) | 1.6E-01 |
| Past smokers |  | 0.203 | (0.255) | 4.3E-01 |
| Current smokers |  | -0.070 | (0.260) | 7.9E-01 |
| Chest RT |  | 1.281 | (0.200) | 1.4E-10 |
| Race |  | 0.255 | (0.315) | 4.2E-01 |
| ***Basic model*** | Pulmonary diffusion deficits |  |  |  |
| (Intercept) |  | -1.517 | (0.450) | 7.5E-04 |
| Attained age, years |  | -0.018 | (0.010) | 6.4E-02 |
| Gender, female |  | 0.175 | (0.160) | 2.7E-01 |
| BMI, <18.5 |  | 1.264 | (0.385) | 1.0E-03 |
| BMI, <18.5-<25.0 |  | 0.500 | (0.199) | 1.2E-02 |
| BMI, ≥25.0 |  | 0.259 | (0.205) | 2.1E-01 |
| Past smokers |  | 0.212 | (0.215) | 3.3E-01 |
| Current smokers |  | 0.757 | (0.182) | 3.2E-05 |
| Chest RT |  | 1.408 | (0.157) | 3.7E-19 |
| Race |  | -0.513 | (0.226) | 2.3E-02 |
| ***Full model additionally adjusted for SDOH*** | Pulmonary diffusion deficits |  |  |  |
| (Intercept) |  | -1.196 | (0.671) | 7.5E-02 |
| Attained age, years |  | -0.010 | (0.011) | 3.5E-01 |
| Gender, female |  | 0.060 | (0.185) | 7.5E-01 |
| Educational attainment |  | -0.104 | (0.092) | 2.6E-01 |
| Personal income |  | -0.156 | (0.084) | 6.5E-02 |
| Area deprivation index, national rank |  | 0.002 | (0.004) | 5.4E-01 |
| BMI, <18.5 |  | 1.364 | (0.421) | 1.2E-03 |
| BMI, <18.5-<25.0 |  | 0.618 | (0.226) | 6.4E-03 |
| BMI, ≥25.0 |  | 0.292 | (0.230) | 2.0E-01 |
| Past smokers |  | 0.159 | (0.239) | 5.0E-01 |
| Current smokers |  | 0.389 | (0.229) | 9.0E-02 |
| Chest RT |  | 1.342 | (0.175) | 1.8E-14 |
| Race |  | -0.344 | (0.256) | 1.8E-01 |
| ***Basic model*** | Restrictive pulmonary deficit |  |  |  |
| (Intercept) |  | -2.294 | (0.570) | 5.7E-05 |
| Attained age, years |  | -0.001 | (0.013) | 9.4E-01 |
| Gender, female |  | -0.106 | (0.212) | 6.2E-01 |
| BMI, <18.5 |  | 0.654 | (0.475) | 1.7E-01 |
| BMI, <18.5-<25.0 |  | -0.133 | (0.262) | 6.1E-01 |
| BMI, ≥25.0 |  | -0.191 | (0.264) | 4.7E-01 |
| Past smokers |  | 0.172 | (0.271) | 5.2E-01 |
| Current smokers |  | -0.284 | (0.280) | 3.1E-01 |
| Chest RT |  | 1.493 | (0.220) | 1.2E-11 |
| Race |  | -0.831 | (0.272) | 2.2E-03 |
| ***Full model additionally adjusted for SDOH*** | Restrictive pulmonary deficit |  |  |  |
| (Intercept) |  | -2.112 | (0.883) | 1.7E-02 |
| Attained age, years |  | -0.001 | (0.015) | 9.3E-01 |
| Gender, female |  | 0.047 | (0.252) | 8.5E-01 |
| Educational attainment |  | -0.191 | (0.122) | 1.2E-01 |
| Personal income |  | -0.041 | (0.113) | 7.1E-01 |
| Area deprivation index, national rank |  | 0.003 | (0.005) | 5.3E-01 |
| BMI, <18.5 |  | 0.760 | (0.529) | 1.5E-01 |
| BMI, <18.5-<25.0 |  | 0.060 | (0.306) | 8.4E-01 |
| BMI, ≥25.0 |  | -0.100 | (0.308) | 7.4E-01 |
| Past smokers |  | 0.367 | (0.300) | 2.2E-01 |
| Current smokers |  | -0.416 | (0.343) | 2.2E-01 |
| Chest RT |  | 1.680 | (0.260) | 1.0E-10 |
| Race |  | -0.830 | (0.318) | 9.0E-03 |
| Abbreviations: SE (standard error), social determinants of health (SDOH), BMI (body mass index), RT (radiation therapy).  *Race was coded as 1 for European American survivors and 0 for African American survivors | | | | |

| **Supplementary Table 4. Association of SDOH associated CpGs with risk of pulmonary diffusion deficits in survivors of European ancestry (*P*_FDR_<0.05)** | | | | | | | |
| --- | --- | --- | --- | --- | --- | --- | --- |
| CpG label | Chr | Position | HGNC gene | Effect size | (SE) | *P_raw_* | *P_FDR_* |
| ***Educational attainment-associated CpG sites and pulmonary diffusion deficits*** | | | | | | |  |
| cg05575921 | chr5 | 373,262 | *AHRR* | -0.94 | (0.16) | 6.7E-09 | 7.2E-07 |
| cg08064403 | chr3 | 98,521,413 | *CLDND1;CPOX;RP11-227H4.5* | -1.17 | (0.21) | 1.4E-08 | 7.2E-07 |
| cg04180924 | chr3 | 98,553,219 | *CPOX* | -1.33 | (0.24) | 2.5E-08 | 7.2E-07 |
| cg19859270 | chr3 | 98,532,449 | *CPOX;GPR15* | -1.20 | (0.22) | 2.6E-08 | 7.2E-07 |
| cg02657160 | chr3 | 98,592,218 | *CPOX* | -2.13 | (0.39) | 3.9E-08 | 8.7E-07 |
| cg00010201 | chr3 | 98,568,261 | *CPOX* | -1.25 | (0.25) | 4.7E-07 | 8.7E-06 |
| cg26768182 | chr9 | 131,397,291 | *PRRC2B* | -1.56 | (0.32) | 1.3E-06 | 2.1E-05 |
| cg05659611 | chr3 | 98,521,779 | *CLDND1;CPOX;RP11-227H4.5* | -1.55 | (0.32) | 1.7E-06 | 2.3E-05 |
| cg02978227 | chr3 | 98,573,182 | *CPOX* | -1.26 | (0.28) | 5.0E-06 | 6.2E-05 |
| cg21566642 | chr2 | 232,419,950 | *ECEL1P1* | -2.12 | (0.48) | 8.8E-06 | 9.8E-05 |
| cg04885881 | chr1 | 11,063,060 | *NA* | -2.10 | (0.48) | 1.3E-05 | 1.3E-04 |
| cg11205006 | chr22 | 26,479,532 | *HPS4* | -1.68 | (0.41) | 4.4E-05 | 4.1E-04 |
| cg00385142 | chr3 | 98,517,073 | *CLDND1* | -1.75 | (0.46) | 1.3E-04 | 8.9E-04 |
| cg13345299 | chr22 | 26,480,108 | *HPS4* | -2.28 | (0.62) | 2.2E-04 | 1.3E-03 |
| cg21431832 | chr22 | 26,479,685 | *HPS4* | -1.66 | (0.45) | 2.4E-04 | 1.3E-03 |
| cg06362176 | chr9 | 124,286,348 | *NEK6* | -2.13 | (0.58) | 2.5E-04 | 1.3E-03 |
| cg03028088 | chr10 | 62,083,461 | *ARID5B* | -2.34 | (0.66) | 4.3E-04 | 2.2E-03 |
| cg00526275 | chr12 | 118,373,745 | *TAOK3* | -1.32 | (0.38) | 5.8E-04 | 2.7E-03 |
| cg02532700 | chr22 | 36,861,361 | *NCF4;NCF4-AS1* | -1.29 | (0.39) | 8.5E-04 | 3.6E-03 |
| cg07178945 | chr12 | 4,379,633 | *FGF23* | 1.36 | (0.43) | 1.7E-03 | 6.9E-03 |
| cg13518852 | chr1 | 212,718,663 | *NA* | 1.02 | (0.36) | 4.8E-03 | 1.8E-02 |
| cg27251814 | chr22 | 26,479,891 | *HPS4* | -1.18 | (0.43) | 5.8E-03 | 1.9E-02 |
| cg12722937 | chr15 | 90,872,816 | *FURIN* | -1.93 | (0.71) | 6.5E-03 | 2.1E-02 |
| cg04791659 | chr15 | 90,872,581 | *FURIN* | -1.16 | (0.43) | 7.4E-03 | 2.3E-02 |
| cg06359375 | chr22 | 26,479,837 | *HPS4* | -1.27 | (0.49) | 9.6E-03 | 3.0E-02 |
| cg15241876 | chr22 | 36,861,551 | *NCF4;NCF4-AS1* | -1.37 | (0.53) | 9.9E-03 | 3.0E-02 |
| cg10698959 | chr15 | 90,872,829 | *FURIN* | -1.66 | (0.67) | 1.4E-02 | 3.9E-02 |
| cg27470486 | chr17 | 41,917,434 | *ACLY* | -1.75 | (0.72) | 1.5E-02 | 4.1E-02 |
| cg03660377 | chr16 | 318,047 | *AXIN1* | -1.17 | (0.49) | 1.8E-02 |  |
| ***Personal income-associated CpG sites and pulmonary diffusion deficits*** | | | | | | |  |
| cg05575921 | chr5 | 373,262 | *AHRR* | -0.94 | (0.16) | 6.7E-09 | 7.2E-07 |
| cg08064403 | chr3 | 98,521,413 | *CLDND1;CPOX;RP11-227H4.5* | -1.17 | (0.21) | 1.4E-08 | 7.2E-07 |
| cg04180924 | chr3 | 98,553,219 | *CPOX* | -1.33 | (0.24) | 2.5E-08 | 7.2E-07 |
| cg19859270 | chr3 | 98,532,449 | *CPOX;GPR15* | -1.20 | (0.22) | 2.6E-08 | 7.2E-07 |
| cg02657160 | chr3 | 98,592,218 | *CPOX* | -2.13 | (0.39) | 3.9E-08 | 8.7E-07 |
| cg00010201 | chr3 | 98,568,261 | *CPOX* | -1.25 | (0.25) | 4.7E-07 | 8.7E-06 |
| cg05659611 | chr3 | 98,521,779 | *CLDND1;CPOX;RP11-227H4.5* | -1.55 | (0.32) | 1.7E-06 | 2.3E-05 |
| cg02978227 | chr3 | 98,573,182 | *CPOX* | -1.26 | (0.28) | 5.0E-06 | 6.2E-05 |
| cg04885881 | chr1 | 11,063,060 | *NA* | -2.10 | (0.48) | 1.3E-05 | 1.3E-04 |
| cg01940273 | chr2 | 232,420,223 | *ECEL1P1* | -1.41 | (0.36) | 7.8E-05 | 6.1E-04 |
| cg06035956 | chr5 | 378,983 | *AHRR* | -1.59 | (0.41) | 8.9E-05 | 6.6E-04 |
| cg00385142 | chr3 | 98,517,073 | *CLDND1* | -1.75 | (0.46) | 1.3E-04 | 8.9E-04 |
| cg18754985 | chr3 | 98,518,905 | *CLDND1* | -1.28 | (0.34) | 1.8E-04 | 1.2E-03 |
| cg27521648 | chr3 | 98,574,171 | *CPOX* | -1.45 | (0.42) | 5.9E-04 | 2.7E-03 |
| cg06235438 | chr16 | 30,474,644 | *ITGAL* | -1.51 | (0.52) | 3.8E-03 | 1.5E-02 |
| cg09422787 | chr14 | 73,741,152 | *ELMSAN1* | -2.04 | (0.84) | 1.5E-02 | 4.1E-02 |
| ***Area deprivation index-associated CpG sites and pulmonary diffusion deficits*** | | | | | | |  |
| cg05575921 | chr5 | 373,262 | *AHRR* | -0.94 | (0.16) | 6.7E-09 | 7.2E-07 |
| cg08064403 | chr3 | 98,521,413 | *CLDND1;CPOX;RP11-227H4.5* | -1.17 | (0.21) | 1.4E-08 | 7.2E-07 |
| cg04180924 | chr3 | 98,553,219 | *CPOX* | -1.33 | (0.24) | 2.5E-08 | 7.2E-07 |
| cg19859270 | chr3 | 98,532,449 | *CPOX;GPR15* | -1.20 | (0.22) | 2.6E-08 | 7.2E-07 |
| cg02657160 | chr3 | 98,592,218 | *CPOX* | -2.13 | (0.39) | 3.9E-08 | 8.7E-07 |
| cg05659611 | chr3 | 98,521,779 | *CLDND1;CPOX;RP11-227H4.5* | -1.55 | (0.32) | 1.7E-06 | 2.3E-05 |
| cg02978227 | chr3 | 98,573,182 | *CPOX* | -1.26 | (0.28) | 5.0E-06 | 6.2E-05 |
| cg08840017 | chr2 | 230,944,894 | *GPR55* | -1.59 | (0.39) | 4.8E-05 | 4.1E-04 |
| cg00385142 | chr3 | 98,517,073 | *CLDND1* | -1.75 | (0.46) | 1.3E-04 | 8.9E-04 |
| cg01899089 | chr5 | 369,853 | *AHRR* | -2.48 | (0.66) | 1.9E-04 | 1.2E-03 |
| cg12803068 | chr7 | 44,963,319 | *MYO1G* | 0.61 | (0.18) | 6.2E-04 | 2.8E-03 |
| cg17585440 | chr19 | 2,052,181 | *MKNK2* | -3.01 | (0.92) | 1.1E-03 | 4.3E-03 |
| cg05221370 | chr7 | 111,098,779 | *IMMP2L;LRRN3* | -0.94 | (0.33) | 5.0E-03 | 1.8E-02 |
| cg26577523 | chr2 | 70,043,693 | *PCBP1-AS1* | -1.65 | (0.60) | 5.8E-03 | 1.9E-02 |
| cg05445320 | chr20 | 47,356,991 | *ZMYND8* | -1.12 | (0.43) | 1.0E-02 | 3.0E-02 |
| Abbreviations: Chr (chromosome), HGNC (The HUGO genome nomenclature committee), SE (standard error), FDR (false-discovery rate), PDD (pulmonary diffusion deficits) | | | | | | | |
